# Decoding WNT Pathway Dysregulation in Bevacizumab-Treated Early-Onset Colorectal Cancer to Inform Precision Oncology

**DOI:** 10.64898/2025.12.22.25342868

**Authors:** Erika Ruiz-Garcia, Brigette Waldrup, Francisco G. Carranza, Sophia Manjarrez, Edith A. Fernandez-Figueroa, Enrique Velazquez-Villarreal

## Abstract

Early-onset colorectal cancer (EOCRC) is rising most rapidly among Hispanic/Latino (H/L) populations, yet the prognostic relevance of WNT pathway alterations under contemporary therapies remains unclear. We conducted a multi-cohort analysis integrating somatic genomics with clinical and treatment annotations (including bevacizumab exposure) from public CRC datasets, stratifying by age of onset and ancestry (H/L vs. non-Hispanic White [NHW]). WNT alterations, dominated by APC, were ubiquitous, but their distribution and outcome associations were context dependent. Across several strata, bevacizumab exposure was associated with lower observed mutation frequencies in select WNT genes (notably RNF43, AXIN1/2, TCF7L2, AMER1), consistent with biological interplay or treatment-related selection. Prognostically, WNT alterations predicted improved overall survival in H/L EOCRC and in NHW late-onset CRC (LOCRC), but worse survival in NHW EOCRC. These findings nominate WNT pathway context as a candidate biomarker for disparity-aware risk stratification in EOCRC and motivate mechanistic studies of anti-angiogenic WNT interactions. Prospective, multi-institutional validation incorporating treatment timing, tumor microenvironment, and social context is warranted to translate these signals into equitable precision oncology.

## 1. Introduction

Colorectal cancer (CRC) remains a significant global health challenge, ranking as the third most commonly diagnosed malignancy and the second leading cause of cancer-related mortality worldwide (1). Despite improvements in screening and treatment in high-income countries, an alarming and persistent rise in CRC among younger adults has emerged. Early-onset CRC (EOCRC), defined as CRC diagnosed before age 50, has increased by approximately 1–2% per year (2, 3). In the late 1990s, CRC was the fourth leading cause of cancer-related death among individuals under 50; today, EOCRC has become the leading cause of cancer death for men and the second for women in this age group (2, 3). Because screening programs generally begin at age 50, EOCRC is frequently detected at more advanced stages, contributing to poorer outcomes.

These trends are further magnified when examined through the lens of racial and ethnic disparities. Although Non-Hispanic Whites (NHWs) currently exhibit the highest absolute incidence of CRC in the United States, Hispanic/Latino (H/L) populations experience the most rapid increase in EOCRC, with annual incidence rising by 2.35% (4–7). Yet, genomic studies have largely focused on NHW populations, creating substantial knowledge gaps regarding EOCRC biology in H/L patients. This is particularly concerning given evidence that H/L individuals derive less benefit from standard CRC therapies and experience poorer overall treatment outcomes compared with NHWs (8). Understanding the biological and genomic drivers of EOCRC in H/L populations is critical to developing equitable precision oncology strategies.

Existing evidence suggests that EOCRC may represent a biologically distinct entity. Several studies report higher microsatellite instability, elevated tumor mutation burden, increased PD-L1 expression, and a greater frequency of hereditary CRC syndromes such as familial adenomatous polyposis in EOCRC (9–13). Others, however, describe differing molecular features, highlighting inconsistencies that warrant deeper investigation. Furthermore, reduced LINE-1 methylation has been proposed as a distinguishing biomarker of EOCRC (14). Growing genomic analyses have begun to explore mutational differences across major CRC signaling pathways, including TP53, KRAS, WNT, and TGF-β, particularly in underserved populations such as H/L patients (15–19). These studies consistently point toward meaningful ancestry-specific variation in tumor biology.

Among the pathways implicated in CRC initiation and progression, WNT signaling plays a central and well-characterized role. WNT pathway activation begins when WNT ligands bind Frizzled receptors and LRP5/6 co-receptors, triggering β-catenin stabilization and downstream transcriptional signaling that drives cell growth and proliferation (20–22). Mutations in key WNT components, particularly the tumor suppressor APC, occur in approximately 80% of CRCs (23). However, studies comparing microsatellite-stable (MSS) EOCRC to later-onset cases suggest that EOCRC may harbor fewer WNT-pathway mutations (13), pointing toward potential differences in pathway activation and therapeutic vulnerabilities in younger patients.

Clinically, EOCRC is more often diagnosed at advanced stages and is associated with increased risk of metastasis and poorer outcomes compared with late-onset CRC (LOCRC) (24, 25). Bevacizumab, a monoclonal antibody targeting vascular endothelial growth factor A (VEGF-A), is a widely used first-line therapy for metastatic CRC (26–28). Its anti-angiogenic mechanism normalizes tumor vasculature, reduces VEGF-driven signaling, and enhances chemotherapy delivery (30, 31). When combined with regimens such as FOLFOX or FOLFIRI, bevacizumab significantly improves progression-free and overall survival, reinforcing its central role in CRC management (30–32). Importantly, bevacizumab’s effectiveness may differ by age and ancestry: younger patients and individuals from underrepresented racial/ethnic groups, including H/L populations, demonstrate variable benefit and distinct toxicity profiles (33, 34).

Emerging data suggest potential biological interactions between WNT and VEGF pathways. Preclinical studies indicate that WNT activation can influence endothelial remodeling, angiogenic signaling, and therapy resistance (35, 36). Alterations in WNT pathway genes, including APC, RNF43, and CTNNB1, have been implicated in modulating tumor responsiveness to anti-angiogenic therapies such as bevacizumab (29). Despite these mechanistic insights, no clinical study has systematically examined how WNT pathway alterations influence bevacizumab outcomes in EOCRC, particularly among H/L patients who are disproportionately affected and historically underrepresented in genomic datasets.

Given the rising incidence of EOCRC in H/L populations, the fundamental role of WNT dysregulation in CRC biology, and the widespread clinical use of bevacizumab, a detailed investigation of WNT pathway alterations in bevacizumab-treated EOCRC across ancestries is urgently needed. In this study, we integrate multi-cohort clinical, genomic, and treatment datasets to characterize WNT pathway alterations in EOCRC, evaluate their association with bevacizumab exposure, and identify ancestry-specific molecular signatures. By leveraging AI-driven precision oncology tools capable of harmonizing demographic, genomic, and therapeutic variables, this work aims to fill critical gaps in EOCRC research and contribute to more equitable, biologically informed treatment approaches for diverse patient populations.

## 2. Materials and Methods

### 2.1. Data Sources and Cohort Assembly

Clinical, genomic, and treatment data for colorectal cancer (CRC) patients were aggregated from multiple publicly accessible datasets hosted within cBioPortal for Cancer Genomics. Datasets were selected based on their availability of comprehensive clinical annotations, chemotherapeutic exposure fields, and high-quality somatic mutation profiles. The final analytic cohort included CRC cases from: TCGA Colorectal Adenocarcinoma (PanCancer Atlas), MSK-CHORD Colorectal Cancer Dataset, and AACR GENIE Biopharma Collaborative (BPC) CRC Cohort.

Only primary adenocarcinomas of the colon and rectum with matched clinical and genomic data were included. For patients with multiple tumor samples, a single primary tumor specimen per individual was retained to avoid duplication bias.

Race and ethnicity were extracted from structured clinical annotations. Individuals labeled as “Hispanic or Latino,” “Hispanic, NOS,” “Spanish, NOS,” or “Latino, NOS” were categorized as Hispanic/Latino (H/L). NHW status was defined as “White” or “Non-Hispanic White.” Age at diagnosis was used to classify patients as EOCRC (<50 years) or LOCRC (≥50 years). Bevacizumab exposure was identified through anticancer drug administration fields and verified manually through treatment timelines when available.

Across all datasets, patients were stratified into subgroups defined by age (EOCRC vs. LOCRC), ancestry (H/L vs. NHW), presence or absence of bevacizumab treatment, and WNT pathway alteration status.

### 2.2. Genomic Annotation and WNT Pathway Classification

Somatic mutation data were retrieved from cBioPortal’s standardized MAF files and converted into gene-level alteration matrices. Mutations were limited to nonsynonymous changes, including missense, nonsense, frameshift insertions/deletions, splice site variants, and translation start site alterations.

WNT pathway genes were curated from established CRC literature and included core members and regulators such as APC, CTNNB1, RNF43, AXIN1, AXIN2, TCF7L2, AMER1, and others. A case was classified as “WNT-altered” if at least one qualifying nonsynonymous mutation was detected in any pathway gene. Gene-level and pathway-level mutation rates were quantified across all demographic and treatment subgroups. Co-alteration patterns, including concurrent mutations within or across pathways, were examined to identify biologically relevant patterns associated with treatment exposure.

### 2.3. Statistical and Survival Analyses

Mutation frequencies between groups were compared using chi-square tests; Fisher’s exact tests were applied when expected cell counts were <5. Comparisons included: EOCRC vs. LOCRC, H/L vs. NHW, bevacizumab-exposed vs. non-exposed, and WNT-altered vs. WNT-wild-type tumors.

Kaplan–Meier survival analyses were performed to assess associations between WNT alterations and overall survival across age, ancestry, and treatment subgroups. Log-rank tests were used to compare survival curves, and median survival with 95% confidence intervals (CIs) was reported. Subgroup analyses were conducted to evaluate whether prognostic effects of WNT alterations differed by bevacizumab exposure.

All statistical analyses were conducted using R (v4.3) and Python (v3.11). A two-sided p-value <0.05 was considered statistically significant.

## 3. Results

### 3.1 Comparison of Clinical and Demographic Characteristics in Hispanic/Latino vs. Non-Hispanic White CRC Cohorts

A total of 275 H/L patients and 2,242 NHW patients with primary colorectal adenocarcinoma were included across all datasets **(Table 1)**. Age-at-onset patterns differed between groups. Within the H/L cohort, 10.9% (n = 30) were classified as EOCRC treated with bevacizumab, 15.6% (n = 43) as EOCRC without bevacizumab, 33.1% (n = 91) as LOCRC treated with bevacizumab, and 40.4% (n = 111) as LOCRC not exposed to bevacizumab. In contrast, the NHW cohort included 7.0% (n = 172) EOCRC treated with bevacizumab, 15.2% (n = 371) EOCRC without bevacizumab, 23.3% (n = 569) LOCRC treated with bevacizumab, and 54.5% (n = 1,330) LOCRC not exposed to bevacizumab. These distributions demonstrate a higher proportional burden of EOCRC among H/L patients and a relatively greater reliance on bevacizumab-containing regimens among LOCRC cases in both populations.

**Table 1.**
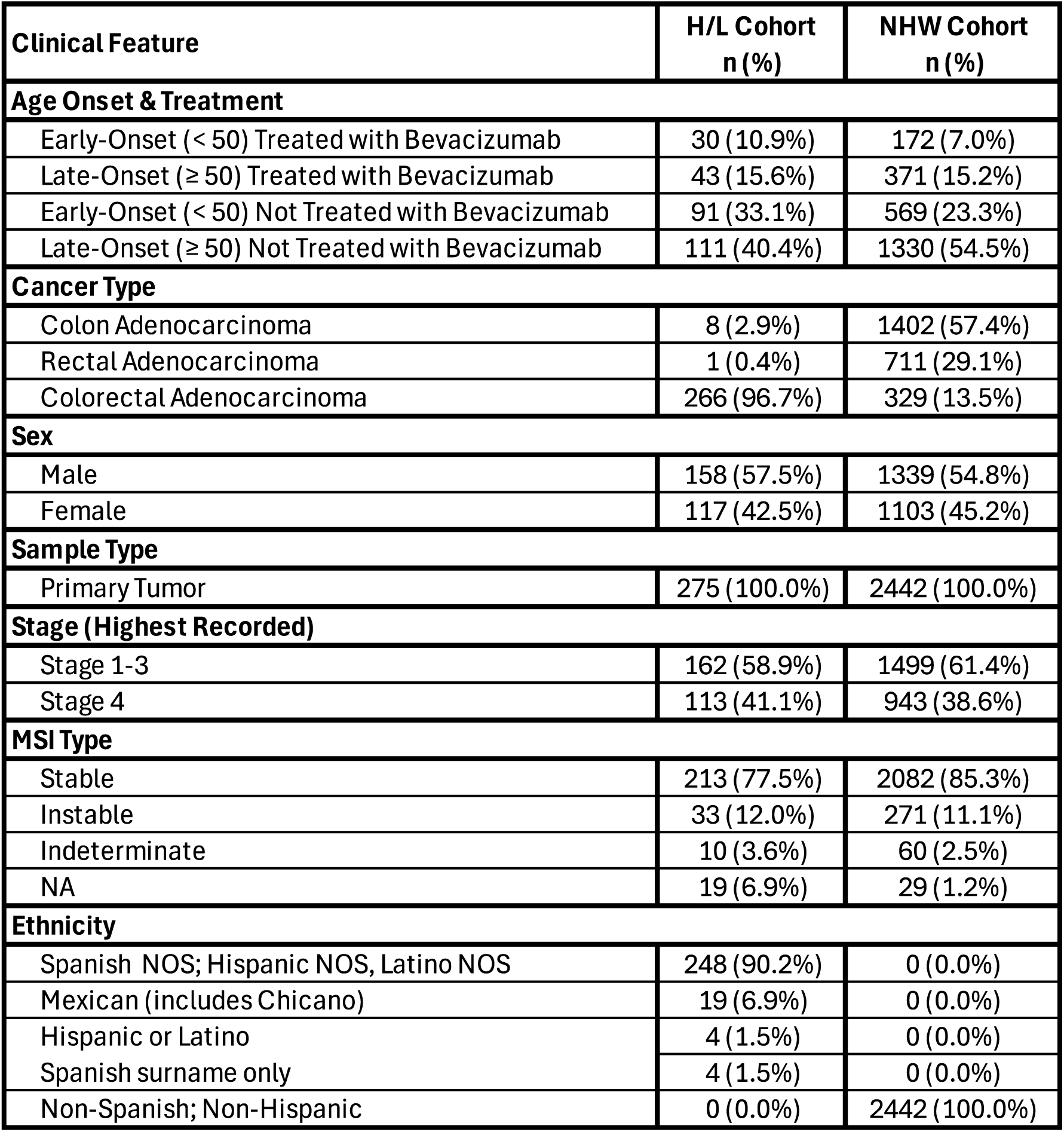
Overview of Clinical Features, Demographics, and Treatment Exposure in H/L and NHW Colorectal Cancer Cohorts.

| Clinical Feature | H/L Cohort<br>n (%) | NHW Cohort<br>n (%) |
| --- | --- | --- |
| **Age Onset & Treatment** |  |  |
| Early-Onset (< 50) Treated with Bevacizumab | 30 (10.9%) | 172 (7.0%) |
| Late-Onset (≥ 50) Treated with Bevacizumab | 43 (15.6%) | 371 (15.2%) |
| Early-Onset (< 50) Not Treated with Bevacizumab | 91 (33.1%) | 569 (23.3%) |
| Late-Onset (≥ 50) Not Treated with Bevacizumab | 111 (40.4%) | 1330 (54.5%) |
| **Cancer Type** |  |  |
| Colon Adenocarcinoma | 8 (2.9%) | 1402 (57.4%) |
| Rectal Adenocarcinoma | 1 (0.4%) | 711 (29.1%) |
| Colorectal Adenocarcinoma | 266 (96.7%) | 329 (13.5%) |
| **Sex** |  |  |
| Male | 158 (57.5%) | 1339 (54.8%) |
| Female | 117 (42.5%) | 1103 (45.2%) |
| **Sample Type** |  |  |
| Primary Tumor | 275 (100.0%) | 2442 (100.0%) |
| **Stage (Highest Recorded)** |  |  |
| Stage 1-3 | 162 (58.9%) | 1499 (61.4%) |
| Stage 4 | 113 (41.1%) | 943 (38.6%) |
| **MSI Type** |  |  |
| Stable | 213 (77.5%) | 2082 (85.3%) |
| Unstable | 33 (12.0%) | 271 (11.1%) |
| Indeterminate | 10 (3.6%) | 60 (2.5%) |
| NA | 19 (6.9%) | 29 (1.2%) |
| **Ethnicity** |  |  |
| Spanish NOS; Hispanic NOS, Latino NOS | 248 (90.2%) | 0 (0.0%) |
| Mexican (includes Chicano) | 19 (6.9%) | 0 (0.0%) |
| Hispanic or Latino | 4 (1.5%) | 0 (0.0%) |
| Spanish surname only | 4 (1.5%) | 0 (0.0%) |
| Non-Spanish; Non-Hispanic | 0 (0.0%) | 2442 (100.0%) |

Cancer type distributions also differed across ancestries. Nearly all H/L patients (96.7%, n = 266) were diagnosed with colorectal adenocarcinoma not otherwise specified, while colon adenocarcinoma and rectal adenocarcinoma constituted only 2.9% and 0.4% of cases, respectively. In contrast, NHW patients displayed a broader distribution, with colon adenocarcinoma accounting for 57.4% (n = 1402), rectal adenocarcinoma for 29.1% (n = 711), and colorectal adenocarcinoma NOS for 13.5% (n = 329). These differences underscore important variation in diagnostic classification and tumor site representation between cohorts.

Sex distribution was similar between groups, with males comprising 57.5% (n = 158) of the H/L cohort and 54.8% (n = 1339) of the NHW cohort. Females represented 42.5% (n = 117) of H/L patients and 45.2% (n = 1103) of NHW patients. All patients (100%) in both cohorts had primary tumor samples, ensuring consistency in tissue type used for genomic analyses.

Stage at diagnosis demonstrated notable variability. Among H/L patients, 58.9% (n = 162) presented with stage 1–3 disease, while 41.1% (n = 113) had stage 4 disease. In comparison, NHW patients showed similar patterns, with 61.4% (n = 1499) diagnosed with stage 1–3 disease and 38.6% (n = 943) diagnosed with stage 4. A small proportion of cases had unknown stage information in both groups (H/L: 5.8%, NHW: 2.9%).

Treatment exposure within each ancestry group also revealed important contrasts. In the H/L cohort, 77.5% (n = 213) received no bevacizumab, while 12.0% (n = 33) received bevacizumab in combination with other regimens, 3.6% (n = 10) received bevacizumab alone, and 6.9% (n = 19) had missing treatment data. Among NHW patients, 85.3% (n = 2082) received no bevacizumab, 11.1% (n = 271) received bevacizumab-containing regimens, and 2.5% (n = 60) received bevacizumab alone, with 1.2% (n = 29) missing treatment information. Collectively, these data illustrate modest but meaningful ancestry-specific differences in bevacizumab administration patterns.

Ethnicity subclassification within the H/L cohort showed that the majority (90.2%, n = 248) were documented as Spanish NOS, Hispanic NOS, or Latino NOS. An additional 9.6% (n = 19) were identified as Mexican (including Chicano), while smaller proportions were categorized as other Hispanic subgroups. By contrast, all NHW patients (100%) were annotated as non-Hispanic White, ensuring a clearly defined comparative framework.

Taken together, these clinical and demographic characteristics highlight distinct patterns in age at onset, cancer classification, staging, and treatment exposure between H/L and NHW patients. The higher proportion of EOCRC in the H/L group, combined with differences in bevacizumab use and diagnostic subtype, emphasizes the need for ancestry-aware molecular analyses to inform precision oncology and address disparities in EOCRC outcomes.

### 3.2 Comparative genomic analysis by age and ancestry

Analysis of H/L patients revealed distinct molecular and genomic patterns when stratifying by age of onset and bevacizumab exposure (Table 2a). Among EOCRC cases, those treated with bevacizumab were diagnosed at a slightly older median age (43 years; IQR: 40–45) compared with their untreated counterparts (41 years; IQR: 36–46), although this difference was not statistically significant (p = 0.12). In contrast, bevacizumab-treated LOCRC patients showed a median diagnosis age similar to untreated LOCRC patients (60 vs. 61 years, p = 0.11). Notably, bevacizumab exposure was associated with substantially lower mutation burden in EOCRC, where treated tumors carried a median of 6 mutations compared with 8 in untreated tumors (p = 0.003). This pattern was mirrored in tumor mutational burden (TMB), with significantly reduced TMB in bevacizumab-treated EOCRC (median 4.9) compared with untreated EOCRC (median 6.1; p = 0.012). Measures of chromosomal instability showed a similar trend: median fraction genome altered (FGA) was higher in bevacizumab-treated EOCRC (0.24) relative to untreated EOCRC (0.17; p = 0.009), whereas no meaningful FGA differences were observed in LOCRC (p = 0.82). WNT-associated gene alterations also differed by treatment status. RNF43, a key negative regulator of WNT signaling and a known marker of pathway dysregulation, was absent in all bevacizumab-treated EOCRC cases (0.0%) but present in 14.3% of untreated EOCRC tumors (p = 0.037). A similar disparity was observed in LOCRC, where RNF43 mutations appeared infrequently in bevacizumab-treated tumors (2.3%) but substantially more often in untreated LOCRC tumors (18.0%, p = 0.0086).

**Table 2.**
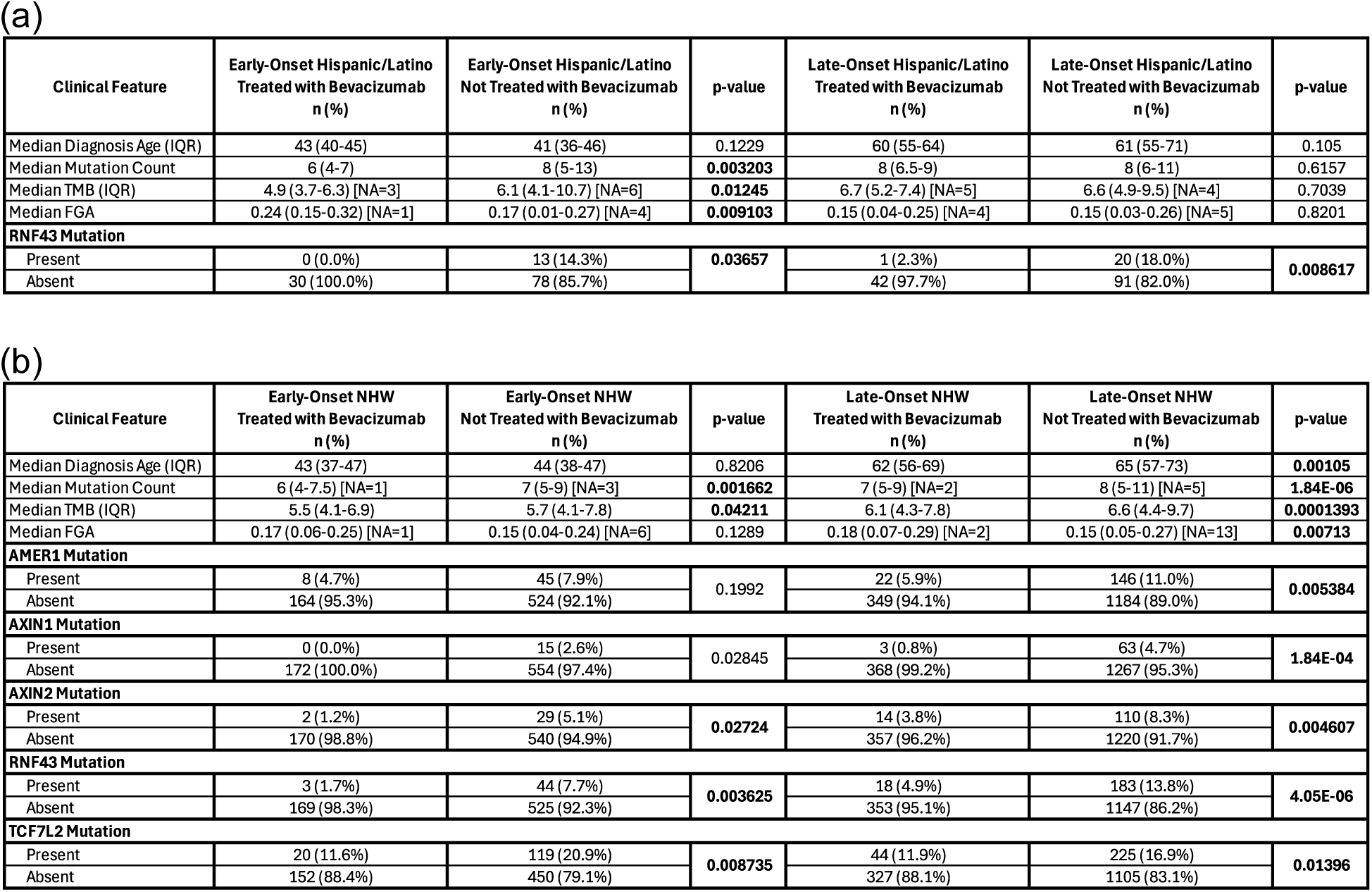
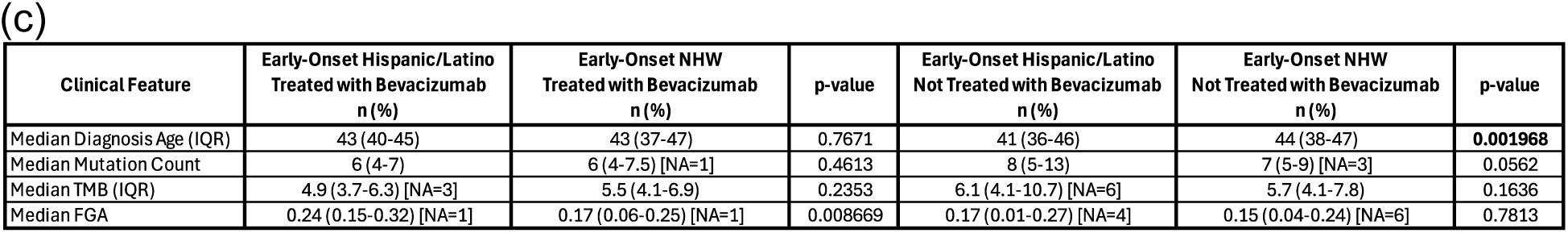
Clinical and genomic comparisons between early-onset and late-onset colorectal cancer (CRC) cohorts. This table presents key clinical and molecular distinctions related to WNT pathway dysregulation and mutation burden across major subgroups: (a) Early-Onset CRC (EOCRC) versus Late-Onset CRC (LOCRC) among Hispanic/Latino (H/L) patients; (b) EOCRC versus LOCRC among Non-Hispanic White (NHW) patients; and (c) cross-ethnic comparisons of EOCRC between H/L and NHW groups. Each analysis includes age at diagnosis, total mutational load, and prevalence of specific WNT pathway gene alterations, organized by ethnicity and age category.

In the NHW cohort, stratification by age of onset and bevacizumab exposure revealed several notable clinical and genomic differences (Table 2b). EOCRC NHW patients showed similar ages at diagnosis regardless of treatment, with median ages of 43 years among bevacizumab-treated individuals and 44 years among untreated patients (p = 0.82). In contrast, LOCRC NHW cases exhibited a modest but statistically significant age difference, with bevacizumab-treated patients diagnosed earlier (median 62 years) than those not exposed to bevacizumab (median 65 years, p = 0.001). Mutation burden differed by treatment status in both age groups: EOCRC tumors receiving bevacizumab carried fewer mutations (median 6) than untreated EOCRC tumors (median 7, p = 0.0017), a pattern that was amplified in LOCRC, where bevacizumab-treated tumors had significantly lower mutation counts than untreated tumors (7 vs. 8, p < 0.0001). Similar reductions were observed for tumor mutational burden (TMB), with bevacizumab-treated cases consistently demonstrating lower TMB values across both EOCRC and LOCRC subsets. Genomic alterations within key WNT pathway regulators also varied by treatment exposure. RNF43 mutations were markedly less frequent among bevacizumab-treated patients in both EOCRC (1.7% vs. 7.7%, p = 0.0036) and LOCRC (4.9% vs. 13.8%, p < 0.00001). A similar pattern was observed for AXIN1 and AXIN2, where bevacizumab-treated tumors exhibited substantially lower mutation rates compared with untreated tumors in both age groups. TCF7L2 mutations followed this trend as well, occurring less often in bevacizumab-treated EOCRC (11.6% vs. 20.9%, p = 0.0087) and LOCRC (11.9% vs. 16.9%, p = 0.014) patients.

Comparison of EOCRC across H/L and NHW patients revealed several ancestry-specific patterns that varied by bevacizumab exposure (Table 2c). Among individuals treated with bevacizumab, H/L and NHW patients exhibited nearly identical ages at diagnosis (median 43 years for both groups, p = 0.77) and comparable mutation burdens, with a median of 6 mutations in each group (p = 0.46). Tumor mutational burden (TMB) also showed no significant ancestry-related differences in the bevacizumab-treated EOCRC subset. However, chromosomal instability differed between groups: H/L patients had significantly higher fraction genome altered (FGA) values than their NHW counterparts (median 0.24 vs. 0.17, p = 0.009), suggesting greater structural genomic disruption in H/L tumors despite similar point mutation loads. In the absence of bevacizumab, age differences between ancestries became more pronounced. Untreated EOCRC H/L patients were diagnosed earlier (median 41 years) than untreated NHW patients (median 44 years, p = 0.002), highlighting a three-year diagnostic gap aligned with known disparities in screening access and symptom recognition. Mutation burden tended to be higher in untreated H/L tumors (median 8) than in untreated NHW tumors (median 7), although this difference approached but did not reach significance (p = 0.056). TMB remained similar between untreated H/L and NHW EOCRC cases, and FGA values showed no meaningful ancestry-related differences in the untreated group.

### 3.3 Distribution of WNT Pathway Alterations by Age, Ethnic Background, and Exposure to Bevacizumab

Across all evaluated subgroups, the integrated analysis revealed that WNT pathway alterations were highly prevalent, with no major differences based on age of onset, ancestral background, or Bevacizumab exposure.

In the H/L cohort, WNT pathway alterations were highly prevalent across both early-and late-onset groups, with little variation by Bevacizumab treatment status (Table 3a). Among early-onset patients, 86.7% of those treated with Bevacizumab exhibited WNT alterations, compared with 91.2% of untreated patients (p = 0.49). A similar pattern was observed in late-onset disease, where WNT alterations were detected in 83.7% of treated cases and 84.7% of untreated cases (p = 1.00). The proportion of patients lacking WNT alterations remained low in all subgroups, ranging from 8.8% to 16.3%. Overall, these results demonstrate that WNT dysregulation is consistently common in H/L CRC, irrespective of age at diagnosis or exposure to Bevacizumab.

**Table 3.** WNT signaling alterations across Hispanic/Latino CRC subgroups classified by age and Bevacizumab therapy. This table presents mutation frequencies for principal WNT pathway genes in H/L CRC patients and includes: (3a) age-of-onset differences within the H/L cohort, (3b) treatment-based variation across EOCRC and LOCRC, (3c) early-onset comparisons between H/L and NHW patients, and (3d) late-onset comparisons by ancestry and treatment status.

| Pathway Alterations | Early-Onset Hispanic/Latino Treated with Bevacizumab n (%) | Early-Onset Hispanic/Latino Not Treated with Bevacizumab n (%) | p-value | Late-Onset Hispanic/Latino Treated with Bevacizumab n (%) | Late-Onset Hispanic/Latino Not Treated with Bevacizumab n (%) | p-value |
| --- | --- | --- | --- | --- | --- | --- |
| WNT Alterations Present | 26 (86.7%) | 83 (91.2%) | 0.4892 | 36 (83.7%) | 94 (84.7%) | 1 |
| WNT Alterations Absent | 4 (13.3%) | 8 (8.8%) |  | 7 (16.3%) | 17 (15.3%) |  |

(b)
| Pathway Alterations | Early-Onset NHW Treated with Bevacizumab n (%) | Early-Onset NHW Not Treated with Bevacizumab n (%) | p-value | Late-Onset NHW Treated with Bevacizumab n (%) | Late-Onset NHW Not Treated with Bevacizumab n (%) | p-value |
| --- | --- | --- | --- | --- | --- | --- |
| WNT Alterations Present | 141 (82.0%) | 499 (87.7%) | 0.07355 | 330 (88.9%) | 1177 (88.5%) | 0.8807 |
| WNT Alterations Absent | 31 (18.0%) | 70 (12.3%) |  | 41 (11.1%) | 153 (11.5%) |  |

(c)
| Pathway Alterations | Early-Onset Hispanic/Latino Treated with Bevacizumab n (%) | Early-Onset NHW Treated with Bevacizumab n (%) | p-value | Early-Onset Hispanic/Latino Not Treated with Bevacizumab n (%) | Early-Onset NHW Not Treated with Bevacizumab n (%) | p-value |
| --- | --- | --- | --- | --- | --- | --- |
| WNT Alterations Present | 26 (86.7%) | 141 (82.0%) | 0.7936 | 83 (91.2%) | 499 (87.7%) | 0.4304 |
| WNT Alterations Absent | 4 (13.3%) | 31 (18.0%) |  | 8 (8.8%) | 70 (12.3%) |  |

(d)
| Pathway Alterations | Late-Onset Hispanic/Latino Treated with Bevacizumab n (%) | Late-Onset NHW Treated with Bevacizumab n (%) | p-value | Late-Onset Hispanic/Latino Not Treated with Bevacizumab n (%) | Late-Onset NHW Not Treated with Bevacizumab n (%) | p-value |
| --- | --- | --- | --- | --- | --- | --- |
| WNT Alterations Present | 36 (83.7%) | 330 (88.9%) | 0.446 | 94 (84.7%) | 1177 (88.5%) | 0.297 |
| WNT Alterations Absent | 7 (16.3%) | 41 (11.1%) |  | 17 (15.3%) | 153 (11.5%) |  |

In the NHW cohort, WNT pathway alterations were also frequent across EOCRC and LOCRC, with only modest differences between patients who received Bevacizumab and those who did not (Table 3b). Among EOCRC cases, 82.0% of treated individuals exhibited WNT alterations compared with 87.7% of untreated individuals (p = 0.0736). In LOCRC, the prevalence of WNT alterations was similarly high and nearly indistinguishable between treated (88.9%) and untreated (88.5%) patients (p = 0.88). Rates of WNT alteration absence remained low, ranging from 11.1% to 18.0% across all subgroups. Collectively, these findings show that WNT dysregulation is widespread in NHW CRC patients, with minimal variation attributable to Bevacizumab exposure or age of onset.

A comparison of EOCRC patients by ancestry showed that WNT pathway alterations were similarly common in both H/L and NHW groups, regardless of Bevacizumab exposure (Table 3c). Among treated patients, 86.7% of H/L individuals exhibited WNT alterations compared with 82.0% of NHW patients (p = 0.79), indicating no meaningful difference between the two groups. In those who did not receive Bevacizumab, WNT alterations were detected in 91.2% of H/L patients and 87.7% of NHW patients (p = 0.43). The proportion of alteration-negative cases remained low across both ancestries, ranging from 8.8% to 18.0%. Overall, WNT dysregulation appears uniformly prevalent in EOCRC irrespective of ancestry or treatment status.

In the LOCRC population, rates of WNT pathway alterations were similarly high among H/L and NHW patients, regardless of treatment status (Table 3d). Among individuals who received Bevacizumab, 83.7% of H/L patients showed WNT alterations compared with 88.9% of NHW patients (p = 0.45). A comparable pattern was observed in untreated patients, with WNT alterations detected in 84.7% of H/L cases and 88.5% of NHW cases (p = 0.30). Across all LOCRC subgroups, the proportion of patients without WNT alterations remained low, ranging from 11.1% to 16.3%. These results indicate that WNT pathway dysregulation is a common hallmark of LOCRC across both ancestry groups, with minimal variation attributable to Bevacizumab exposure.

### 3.4 Frequencies of Gene Alterations in the WNT Pathway

#### Gene-level WNT pathway alterations in early-onset Hispanic/Latino patients by Bevacizumab treatment

To explore whether Bevacizumab exposure was associated with distinct genomic patterns in EOCRC in H/L, we compared mutation frequencies across key WNT pathway genes (Table S1). Overall, most genes showed low mutation rates with no statistically significant differences between treated and untreated groups. APC alterations remained highly prevalent in both Bevacizumab-treated (80.0%) and untreated (83.5%) patients (p = 0.87), consistent with its fundamental role in canonical WNT signaling. Mutations in AMER1, AXIN1, AXIN2, CTNNB1, GSK3B, TCF7L2, TLE1, and TLE2 occurred infrequently and did not differ significantly by treatment status. The exception was RNF43, which showed a notable enrichment in untreated patients (14.3%) compared with no detected mutations in the treated group (0%; p = 0.0366), suggesting a potential treatment-related selection effect on this non-APC regulatory component of the WNT pathway.

#### Gene-Level WNT pathway alterations in late-onset Hispanic/Latino patients by Bevacizumab treatment

In the LOCRC H/L CRC, comparison of gene-level WNT pathway alterations revealed largely similar mutation patterns between patients treated with Bevacizumab and those who were untreated (Table S2). APC remained the most frequently mutated gene in both groups, occurring in 79.1% of treated individuals and 65.8% of untreated individuals, though this difference did not reach statistical significance (p = 0.16). Several genes, including AMER1, CTNNB1, TCF7L2, and AXIN1, displayed low mutation frequencies overall, with no meaningful treatment-associated differences. RNF43 showed the most notable variation, with a markedly higher mutation rate in untreated patients (18.0%) compared with treated patients (2.3%), a statistically significant difference (p = 0.0086), suggesting a potential selective effect or underlying biological distinction in this subgroup.

#### Comparison between early- and late-onset Hispanic/Latino patients treated with Bevacizumab

When evaluating WNT pathway alterations among H/L patients treated with Bevacizumab, EOCRC and LOCRC groups exhibited highly similar mutation profiles (Table S3). APC mutations remained the most common alteration in both EOCRC (80.0%) and LOCRC (79.1%) patients, with no detectable difference between the two groups (p = 1.00). Mutations in AMER1, CTNNB1, TCF7L2, and AXIN2 occurred infrequently and showed no statistically significant variation by age of onset. Likewise, alterations in AXIN1, GSK3B, TLE1, and TLE2 were absent across both cohorts. RNF43 mutations were detected only in the LOCRC group (2.3%), but this difference did not reach significance.

#### Comparison between untreated early- and late-onset Hispanic/Latino patients

In the subset of H/L patients who did not receive Bevacizumab, several WNT pathway genes showed notable age-associated differences (Table S4). APC mutations were significantly more common in EOCRC than in LOCRC (83.5% vs. 65.8%; p = 0.0071), underscoring its central role in EOCRC tumorigenesis. Although most other genes demonstrated similar mutation rates between groups, CTNNB1 and RNF43 showed modest variations, with slightly higher frequencies in EOCRC (9.9% and 14.3%, respectively) compared with LOCRC (6.3% and 18.0%), though these differences were not statistically significant. Mutations in AMER1, AXIN1, AXIN2, TCF7L2, TLE1, and TLE2 occurred infrequently and were comparable across age groups.

#### Gene-level WNT alterations in early-onset Non-Hispanic White patients by Bevacizumab treatment

Among EOCRC NHW patients, evaluation of WNT pathway genes revealed several differences between those treated with Bevacizumab and those who were untreated, although most did not reach statistical significance (Table S5). APC mutations were the most common alteration in both groups, occurring in 75.0% of treated patients and 80.8% of untreated patients (p = 0.12). While mutations in CTNNB1 and GSK3B were rare and comparable across groups, certain genes, including AXIN1, AXIN2, RNF43, and TCF7L2, showed higher mutation frequencies among untreated individuals, with AXIN1 (2.6%), AXIN2 (5.1%), RNF43 (7.7%), and TCF7L2 (20.9%) all elevated relative to treated patients (0–1.7%). Several of these differences were statistically significant, suggesting potential treatment-associated selection patterns. Mutations in AMER1 occurred infrequently and showed no meaningful variation by treatment status.

#### Gene-level WNT alterations in late-onset Non-Hispanic White patients by Bevacizumab treatment

In LOCRC NHW patients, comparison of WNT pathway mutations revealed clear differences between those treated with Bevacizumab and those who were untreated (Table S6). Several non-canonical WNT regulators showed significantly reduced mutation frequencies in the treated group, including AXIN1 (0.8% vs. 4.7%; p = 1.84×10⁻⁴), AXIN2 (3.8% vs. 8.3%; p = 0.0046), RNF43 (4.9% vs. 13.8%; p = 4.05×10⁻⁶), and TCF7L2 (11.9% vs. 16.9%; p = 0.014), suggesting a potential treatment-associated selection against these alterations. AMER1 mutations were also significantly less frequent in treated patients (5.9% vs. 11.0%; p = 0.0054). In contrast, APC mutations, while slightly more common in treated individuals (80.1% vs. 75.1%), did not differ significantly. Mutations in CTNNB1 and GSK3B were infrequent and showed no meaningful variation by treatment status.

#### Comparison between early- and late-onset Non-Hispanic White patients treated with Bevacizumab

Among NHW patients receiving Bevacizumab, the distribution of WNT pathway mutations was largely consistent between EOCRC and LOCRC CRC (Table S7). APC mutations were the most common alteration in both cohorts—75.0% in EOCRC and 80.1% in LOCRC (p = 0.22), reflecting their central role in WNT-driven tumorigenesis. Mutations in other genes, including AMER1, CTNNB1, TCF7L2, and GSK3B, occurred at similarly low frequencies across both age groups. Although RNF43 and AXIN2 mutations appeared slightly more frequent in LOCRC than EOCRC, these differences did not reach statistical significance.

#### Comparison between early- and late-onset Non-Hispanic White patients not treated with Bevacizumab

In untreated NHW patients, several WNT pathway genes displayed significant age-associated differences (Table S8). APC mutations were notably more frequent in EOCRC than in LOCRC (80.8% vs. 75.1%; p = 0.0080), highlighting stronger canonical WNT involvement in younger patients. Conversely, multiple non-canonical regulators, including AXIN1 (4.7% vs. 2.6%; p = 0.047), AXIN2 (8.3% vs. 5.1%; p = 0.019), and RNF43 (13.8% vs. 7.7%; p = 2.82×10⁻⁴), were significantly enriched in the late-onset group, indicating a shift toward alternative WNT signaling mechanisms with increasing age. AMER1 and TCF7L2 mutations also differed modestly by age, with AMER1 more common in LOCRC and TCF7L2 slightly more frequent in EOCRC. CTNNB1 and GSK3B showed no substantial differences.

#### Comparison between early-onset Hispanic/Latino and Non-Hispanic White patients treated with Bevacizumab

Table S9 presents a cross-ancestry comparison of WNT pathway mutations among Bevacizumab-treated EOCRC patients of H/L and NHW backgrounds. Across all evaluated genes, mutation frequencies were highly similar between ancestries, with no statistically significant differences observed. APC mutations, the most common alteration in both groups, occurred at comparable rates (80.0% in H/L vs. 75.0% in NHW), while CTNNB1, AXIN2, TCF7L2, and RNF43 mutations were detected at low but nearly identical frequencies. Several genes, including AXIN1, GSK3B, TLE1, and TLE2, showed no mutations in either group.

#### Comparison between untreated early-onset Hispanic/Latino and Non-Hispanic white patients

Among untreated EOCRC patients (Table S10), several WNT pathway genes showed modest ancestry-related differences, although none reached statistical significance. APC mutations were common in both groups, occurring at similar frequencies in H/L (83.5%) and NHW (80.8%) patients. While CTNNB1 and RNF43 mutations appeared somewhat more frequent in H/L patients (9.9% and 14.3%, respectively) compared with NHW patients (6.0% and 7.7%), these trends did not meet significance thresholds. Mutation rates for AMER1, AXIN1, AXIN2, TCF7L2, and GSK3B were largely comparable across ancestries.

#### Comparison between late-onset Hispanic/Latino and Non-Hispanic White patients treated with Bevacizumab

Among Bevacizumab-treated LOCRC patients (Table S11), gene-level WNT mutation patterns were broadly comparable between H/L and NHW groups. APC remained the most frequently mutated gene in both ancestries, with nearly identical frequencies (79.1% in H/L vs. 80.1% in NHW; p = 1.00). Mutations in AMER1, CTNNB1, AXIN2, and TCF7L2 occurred at low but similar rates across groups. RNF43 alterations were slightly more common in NHW patients (4.9%) compared with H/L patients (2.3%), although this difference was not statistically significant.

#### Comparison between late-onset Hispanic/Latino and Non-Hispanic White patients not treated with Bevacizumab

Among untreated LOCRC patients (Table S12), WNT pathway mutation patterns were broadly similar between H/L and NHW groups, with no statistically significant ancestry-associated differences identified. APC mutations were slightly more frequent in NHW patients (75.1%) than in H/L patients (65.8%; p = 0.040), representing the only gene with a significant difference between groups. Other alterations—including those in AXIN1, AXIN2, CTNNB1, RNF43, and TCF7L2, occurred at comparable rates across ancestries, with AXIN2 mutations showing nearly identical frequencies (7.2% in H/L vs. 8.3% in NHW).

### 3.5 Mutational Landscape

#### Mutational landscape of WNT pathway in early-onset Hispanic/Latino CRC

To define the somatic mutation landscape of WNT signaling in EOCRC H/L patients, we examined the distribution of alteration types and Bevacizumab treatment status using the oncoplot visualization (Figure 1A)), complemented by detailed classification of variant types (Table S13). Nearly all EO H/L tumors exhibited at least one alteration in a WNT pathway gene, underscoring the central role of WNT dysregulation in early-onset carcinogenesis in this population. APC was the most frequently mutated gene, showing a dense pattern of truncating alterations—including nonsense and frameshift events— consistent with its canonical function as the gatekeeping regulator of β-catenin degradation. RNF43, AMER1, CTNNB1, and TCF7L2 represented the next most commonly altered WNT regulators, each displaying a heterogeneous mutation spectrum that included missense, frameshift, and splice-site events, indicative of diverse mechanisms of pathway disruption. Less frequently mutated genes such as AXIN1, AXIN2, GSK3B, and TLE family members appeared only sporadically, with isolated mutations distributed across a subset of EO tumors.

**Figure 1.**
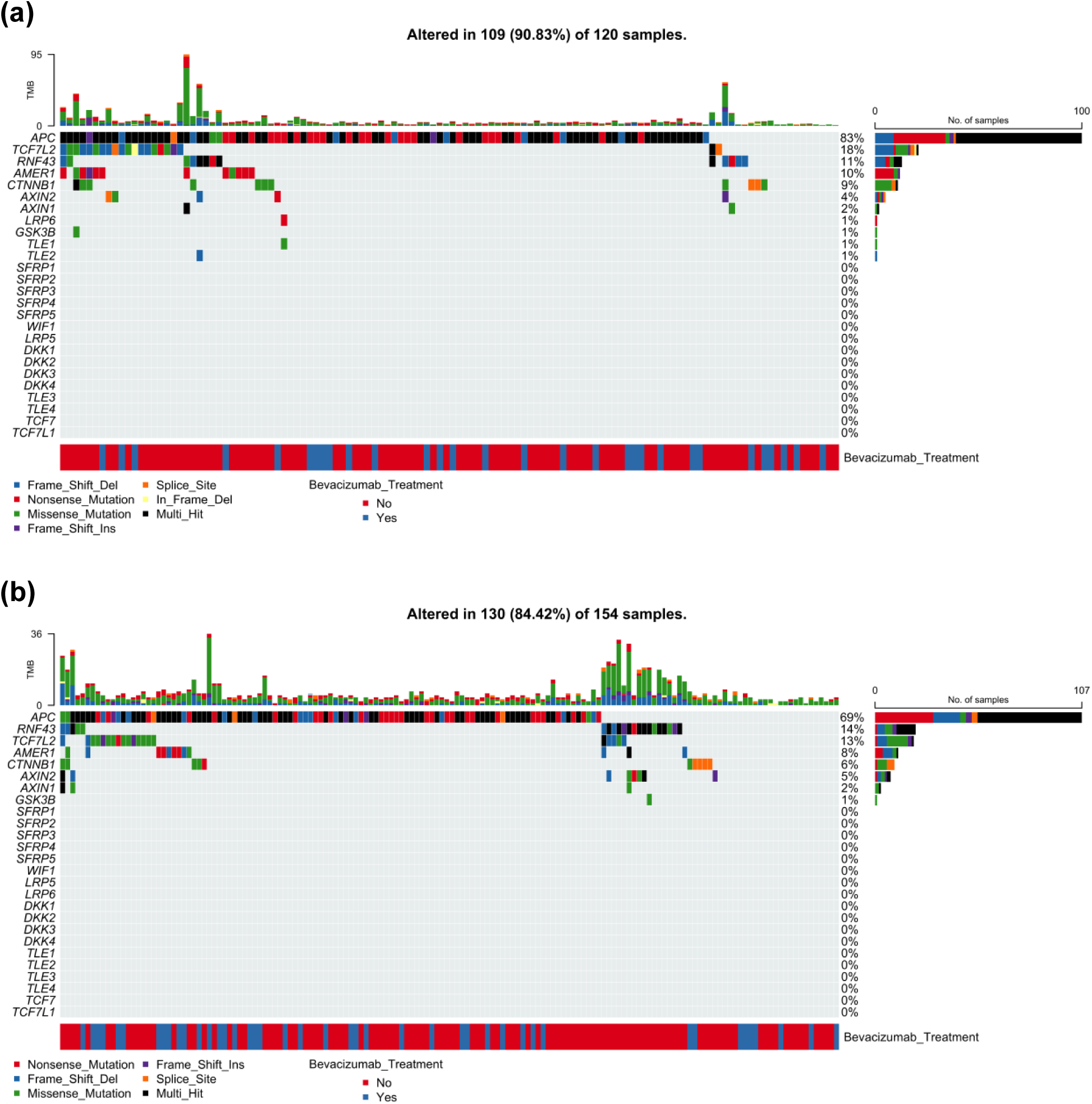
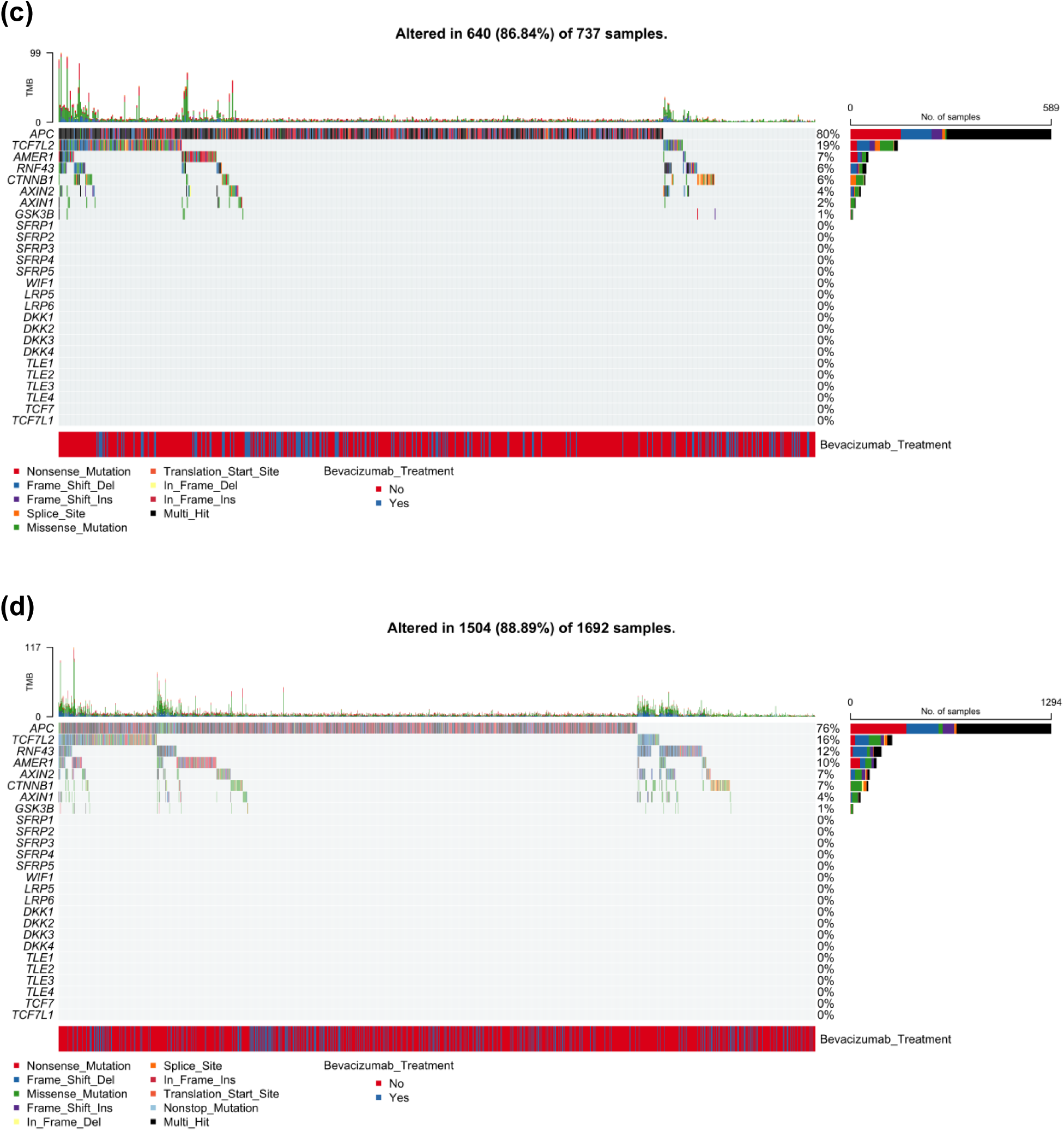
Somatic mutation profiles of WNT pathway genes in colorectal cancer (CRC) stratified by age and ancestry. Oncoplots illustrate gene-level alteration patterns across WNT pathway components in CRC, separated by early-vs. late-onset disease and by H/L vs. NHW ancestry. Each panel displays mutation classes, tumor mutational burden (TMB), and Bevacizumab treatment status for: (a) 120 early-onset H/L patients, (b) 154 late-onset H/L patients, (c) 737 early-onset NHW patients, and (d) 1692 late-onset NHW patients. APC emerges as the most frequently altered gene across all groups, dominated by truncating variants. Additional recurrent mutations in RNF43, TCF7L2, AMER1, and CTNNB1 underscore widespread disruption of WNT signaling, with age- and ancestry-related variation evident in the mutational patterns.

Tumor mutational burden (TMB), shown in the upper panel, was generally low across EO H/L samples, although a small fraction demonstrated elevated TMB values, suggesting potential hypermutated or mismatch-repair–deficient phenotypes within this subgroup. The Bevacizumab annotation track at the bottom of the figure revealed substantial intermixing of treated and untreated cases, with no evident clustering of mutation types or frequencies by treatment exposure.

#### WNT pathway alterations in late-onset Hispanic/Latino CRC

To evaluate the somatic alteration landscape of the WNT signaling pathway in LOCRC among H/L patients, we analyzed mutation patterns and Bevacizumab treatment status using the oncoplot visualization (Figure 1B), supported by mutation classification data (Table S13). Across the 154 LO H/L CRC samples displayed, the majority (84.4%) harbored at least one WNT pathway alteration, demonstrating that WNT dysregulation remains a defining molecular hallmark even in later-onset disease.

APC was the most frequently mutated gene, showing extensive truncating events, primarily nonsense and frameshift mutations, consistent with its established role as a key tumor suppressor governing β-catenin turnover. RNF43 represented the next most commonly altered WNT regulator, exhibiting a heterogeneous distribution of mutation types, including frameshift deletions, splice-site alterations, and multi-hit events. TCF7L2 and AMER1 were also recurrently mutated, showing a mixture of missense, frameshift, and splice-site variants, reflecting diverse mechanisms of WNT pathway disruption within LO H/L tumors. Less frequently mutated genes such as CTNNB1, AXIN1, AXIN2, and GSK3B appeared in a smaller subset of samples, but when present, contributed additional layers of canonical and non-canonical WNT perturbation.

Tumor mutational burden (TMB), illustrated in the top panel of Figure 1B, was generally low across samples, although a subset exhibited moderately elevated TMB values, potentially indicative of mismatch repair deficiency or underlying hypermutation. The Bevacizumab annotation bar at the bottom of the oncoplot revealed substantial intermixing of treated and untreated tumors, with no clear separation in mutation profiles or burden across treatment groups.

#### WNT pathway alterations in Early-Onset Non-Hispanic White CRC

To define the mutational landscape of WNT signaling in EOCRC among NHW patients, we examined somatic alteration patterns and Bevacizumab treatment status using the oncoplot shown in Figure 1C, and mutation classification data in Table S13. Across the 737 EO NHW tumors represented, 640 (86.8%) harbored at least one alteration in a WNT pathway gene, confirming the pervasive involvement of WNT dysregulation in EOCRC within this population.

APC was by far the most frequently mutated gene, with a dense distribution of truncating mutations, including nonsense and frameshift events, consistent with its role as the primary tumor suppressor governing canonical WNT activity. Additional recurrently altered genes included TCF7L2, RNF43, AMER1, and CTNNB1, each showing heterogeneous patterns of mutation types such as missense substitutions, splice-site alterations, in-frame changes, and multi-hit events, indicating multiple mechanisms through which Wnt signaling becomes disrupted. Lower-frequency mutations were observed in AXIN1, AXIN2, GSK3B, and TLE-family genes, typically as isolated missense or frameshift variants scattered across the cohort.

Tumor mutational burden (TMB), depicted in the upper panel, was generally low but exhibited a subset of cases with markedly elevated TMB, suggesting possible hypermutator or mismatch-repair–deficient phenotypes in a minority of EO NHW tumors. The Bevacizumab treatment annotation in the lower panel revealed an intermixing of treated and untreated cases throughout the mutation landscape, with no clear clustering by treatment exposure.

#### WNT pathway alterations in Late-Onset Non-Hispanic White CRC

To define the mutational architecture of the WNT signaling pathway in LOCRC among NHW patients, we examined somatic alteration patterns and Bevacizumab treatment status using oncoplot visualization (Figure 1D) complemented by variant types (Table S13). Across the 1,692 LO NHW tumors analyzed, 1,504 (88.9%) harbored at least one alteration in a WNT pathway gene, confirming that WNT dysregulation remains a fundamental driver of tumorigenesis in older NHW patients.

APC was the most frequently mutated gene, with a dense pattern of truncating events, including nonsense and frameshift mutations, consistent with its longstanding role as the central tumor suppressor controlling β-catenin degradation. Additional recurrent alterations were observed in TCF7L2, RNF43, AMER1, CTNNB1, and AXIN2. Each of these genes displayed a heterogeneous spectrum of mutation types, including missense, frameshift, splice-site, and in-frame variants, reflecting diverse mechanisms of canonical and non-canonical WNT pathway disruption. CTNNB1 alterations were largely composed of missense and in-frame events consistent with potential stabilization of β-catenin, while RNF43 mutations frequently included frameshift and splice-site changes suggestive of loss-of-function. AXIN1, GSK3B, and several TLE family members appeared only in a small minority of samples, typically as isolated missense or frameshift variants.

Tumor mutational burden (TMB), illustrated in the upper panel of Figure 1D, was generally low to moderate across the cohort, though a subset of tumors displayed elevated TMB values, potentially indicative of mismatch repair deficiency or underlying hypermutation. Bevacizumab treatment status, annotated in the lower panel, was intermixed across the mutation landscape, showing no clear clustering or enrichment of mutation types by treatment exposure.

### 3.6 Survival analysis

We evaluated the association between WNT pathway alterations and overall survival across colorectal cancer subgroups defined by age, ancestry, and Bevacizumab treatment status using Kaplan-Meier analysis.

Among EOCRC H/L patients treated with Bevacizumab, overall survival did not differ significantly between individuals with WNT pathway alterations and those without them (p = 0.6; Figure 2a). The survival curves remained closely aligned throughout the follow-up period, with broad and overlapping confidence intervals that indicate substantial uncertainty, particularly within the WNT-unaltered subgroup, which had very limited sample size. While the WNT-altered group showed modest declines in survival probability over time, these fluctuations were not meaningfully distinct from the trajectory of the unaltered group. The small number of WNT-unaltered cases (n=4 at baseline) further limits interpretability. Overall, these findings suggest that, within this Bevacizumab-treated EOCRC H/L cohort, WNT pathway status does not appear to exert a measurable impact on survival outcomes.

**Figure 2.**
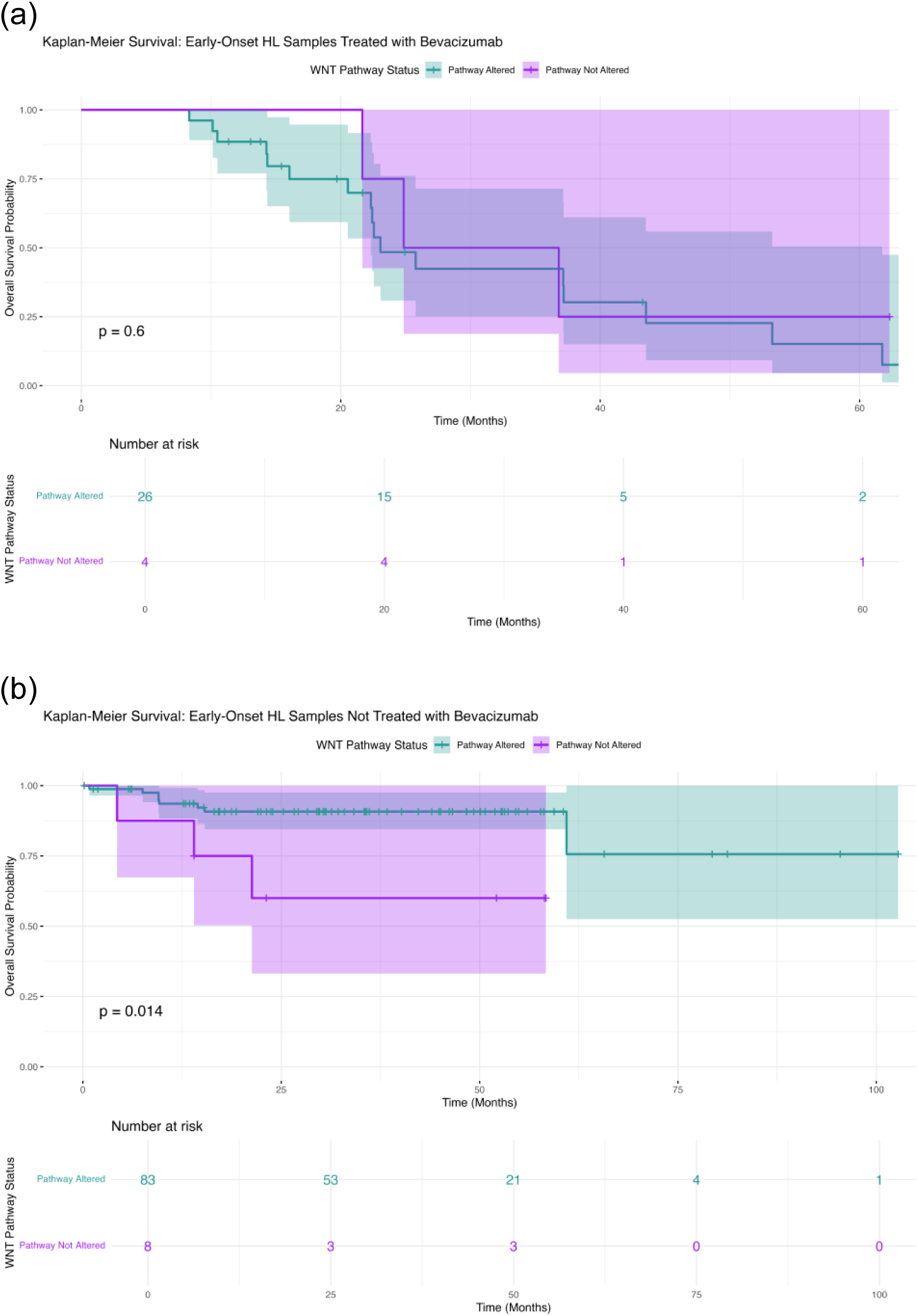
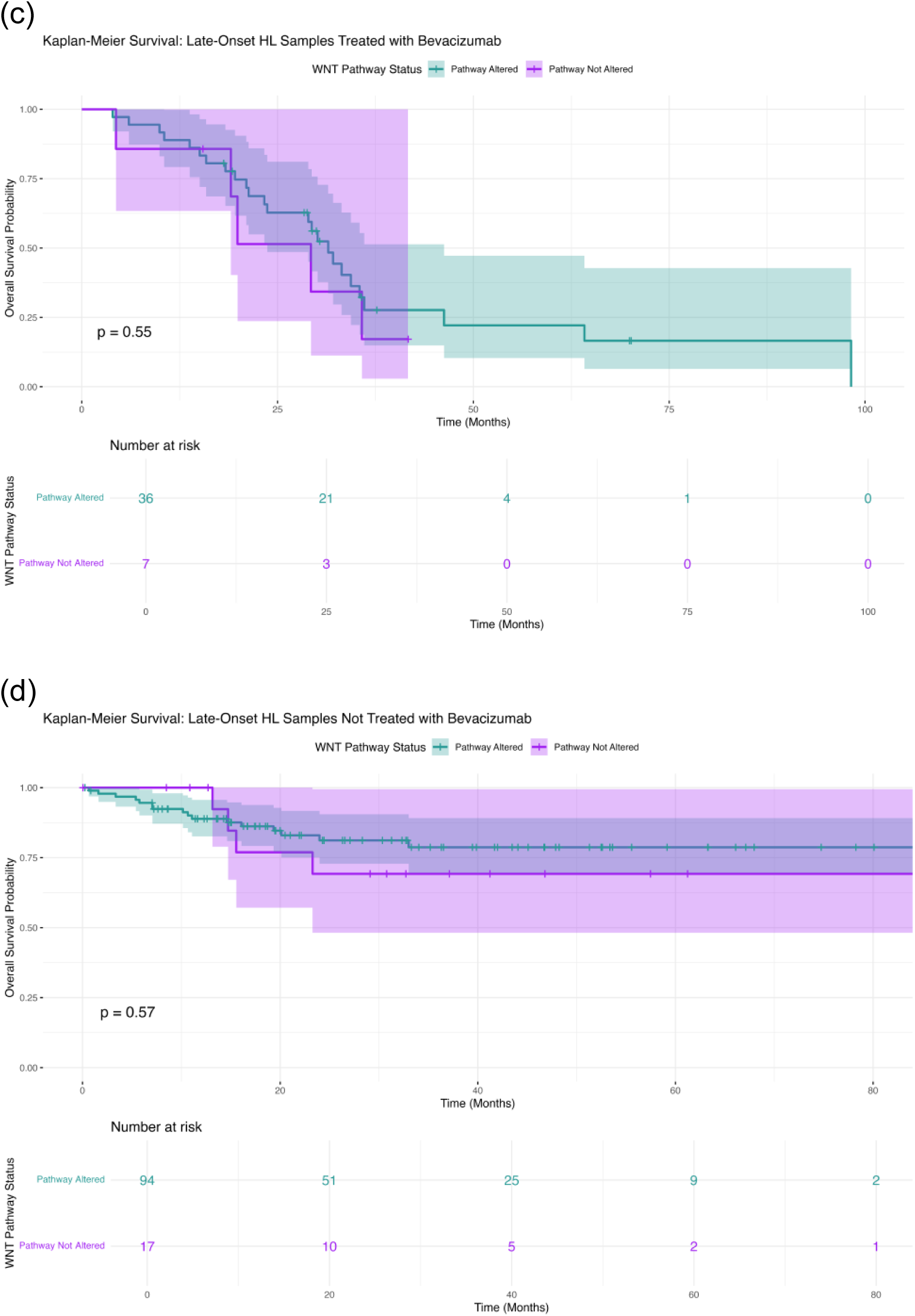
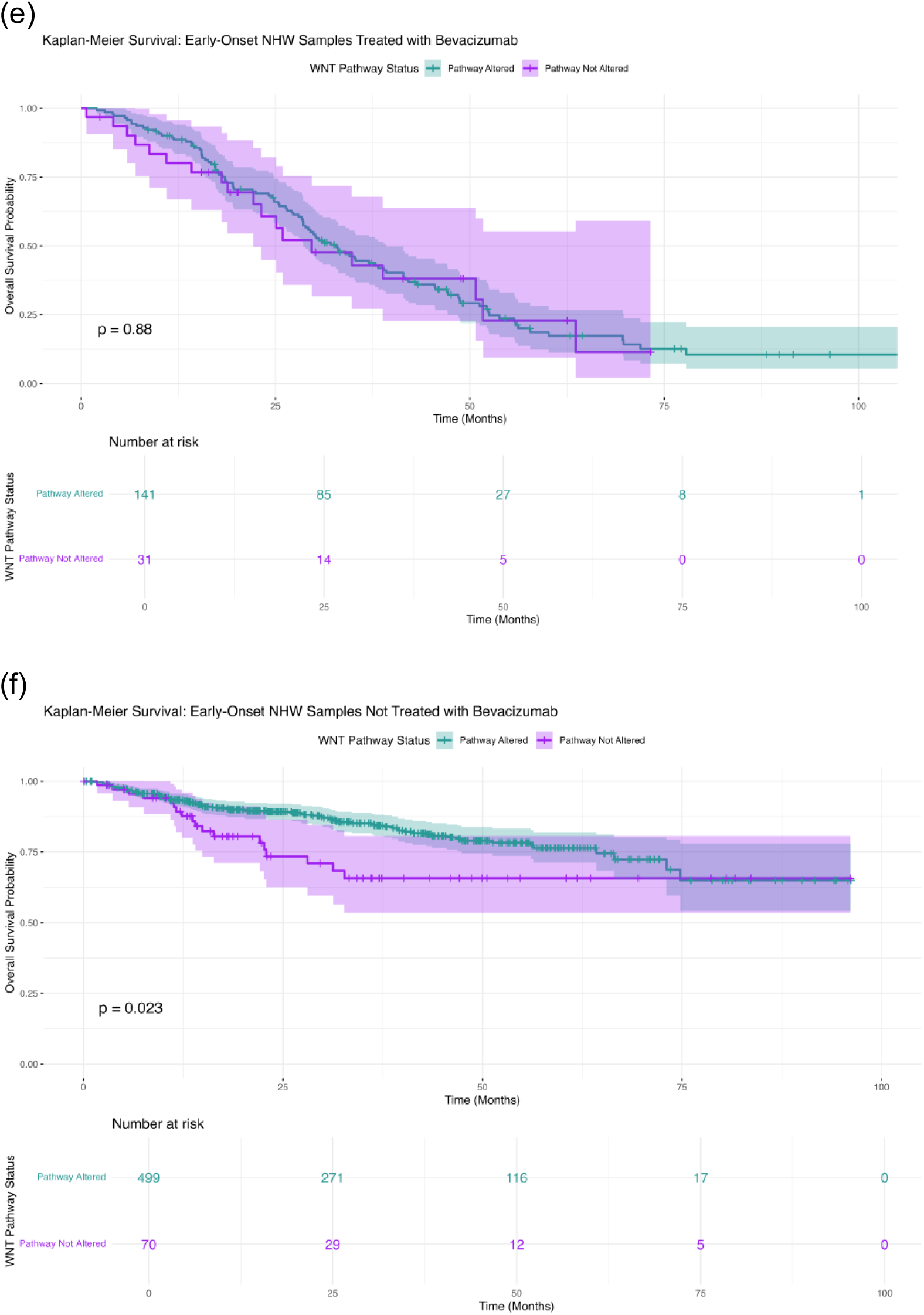
Somatic mutation profiles of WNT pathway genes across colorectal cancer subgroups stratified by age, ancestry, and Bevacizumab exposure. Oncoplots display mutation frequencies and variant types for six groups: (a) Early-Onset Hispanic/Latino (H/L) treated with Bevacizumab, (b) Early-Onset H/L without Bevacizumab, (c) Late-Onset H/L treated with Bevacizumab, (d) Late-Onset H/L without Bevacizumab, (e) Early-Onset Non-Hispanic White (NHW) treated with Bevacizumab, and (f) Early-Onset NHW without Bevacizumab.

In EOCRC H/L patients who did not receive Bevacizumab, overall survival differed significantly by WNT pathway status (p = 0.014; Figure 2b). Patients with WNT pathway alterations demonstrated notably better survival over time, with a stable trajectory extending beyond 100 months, whereas those without WNT alterations exhibited earlier and more pronounced declines in survival probability. The separation between the curves became apparent within the first 40 months of follow-up and persisted thereafter. However, the wide confidence intervals surrounding the WNT-unaltered group, driven by the very small sample size (n = 8 at baseline), introduce uncertainty and limit the robustness of the comparison. Despite this limitation, the observed divergence suggests a potential survival disadvantage associated with the absence of WNT alterations in untreated EO H/L tumors, warranting further investigation in larger cohorts.

LOCRC H/L patients treated with Bevacizumab showed no significant difference in overall survival based on WNT pathway status (p = 0.55; Figure 2c). The Kaplan–Meier curves for WNT-altered and non-altered tumors tracked closely throughout the follow-up period, with broad overlap in their confidence intervals. A gradual decline in survival was observed in both groups between 25 and 75 months, but neither trajectory diverged meaningfully enough to suggest a WNT-dependent survival effect. As the number of patients at risk diminished beyond 60 months, particularly in the unaltered subgroup, the confidence intervals widened substantially, reflecting increased uncertainty in late follow-up estimates. Overall, these data indicate that, among Bevacizumab-treated LOCRC H/L patients, WNT pathway alterations do not appear to influence survival outcomes.

Among LOCRC H/L patients who did not receive Bevacizumab, overall survival did not differ significantly between those with and without WNT pathway alterations (p = 0.57; Figure 2d). The survival curves remained tightly aligned throughout the follow-up period, with only a minor early divergence favoring the unaltered group that quickly converged as time progressed. Confidence intervals widened substantially after approximately 40–50 months, particularly for the WNT-unaltered subgroup, reflecting diminishing numbers at risk and increased uncertainty in long-term estimates. Overall, these results indicate no measurable impact of WNT pathway status on survival outcomes in untreated LOCRC H/L patients.

Among EOCRC NHW patients treated with Bevacizumab, WNT pathway status did not significantly influence overall survival (p = 0.88; Figure 2e). The survival curves for WNT-altered and unaltered groups were nearly superimposed across the entire follow-up period, indicating highly similar survival trajectories. Although minor fluctuations appeared at intermediate time points, these did not translate into meaningful or persistent differences in outcome. Confidence intervals for both groups showed moderate widening after approximately 60 months, consistent with decreasing numbers at risk, but remained broadly overlapping throughout. Overall, these findings suggest that WNT alterations do not confer a survival advantage or disadvantage in EOCRC NHW patients receiving Bevacizumab.

Among EOCRC NHW patients who did not receive Bevacizumab, WNT pathway status was significantly associated with overall survival (p = 0.023; Figure 2f). Patients with WNT pathway alterations demonstrated notably better survival throughout the follow-up period, with the curves separating early and remaining consistently apart over time. In contrast, the WNT-unaltered group exhibited a steeper decline in survival probabilities beginning in the first 20–30 months. Although confidence intervals broadened in both groups at later time points, reflecting fewer individuals at risk, the separation between curves persisted, indicating a meaningful survival advantage for WNT-altered patients in this cohort.

Among LOCRC NHW patients treated with Bevacizumab, WNT pathway status was not associated with a significant difference in overall survival (p = 0.38; Figure S1a). The survival curves for WNT-altered and unaltered tumors closely overlapped across the observation period, indicating comparable outcomes regardless of mutation status. A modest early separation, slightly favoring the WNT-altered group, narrowed quickly as follow-up progressed, with both groups demonstrating similar long-term survival patterns. Confidence intervals expanded substantially after 50 months due to decreasing numbers at risk, limiting the precision of late-survival estimates. Overall, these findings suggest that in Bevacizumab-treated LOCRC NHW patients, WNT pathway alterations do not materially influence survival outcomes.

In H/L NHW patients who did not receive Bevacizumab, WNT pathway alterations were significantly associated with improved overall survival (p = 0.0052; Figure S1b). The survival curves began separating early in the follow-up period, with WNT-altered patients consistently demonstrating higher survival probabilities across nearly the entire time span. This advantage remained evident through mid-and late-follow-up, reflecting a sustained survival benefit in the altered group. Confidence intervals were relatively steady for most of the timeline but expanded past 75–90 months as the number of patients at risk declined, introducing greater uncertainty in long-term estimates. Overall, these results indicate that in untreated LOCRC NHW patients, WNT pathway alterations mark a subgroup with more favorable survival outcomes.

## 4. Discussion

This AI-driven, multi-cohort analysis provides a pathway-level view of WNT dysregulation in colorectal cancer, integrating age of onset, ancestry, and Bevacizumab exposure. By leveraging AI-HOPE-WNT to harmonize clinical, treatment, and genomic features, we were able to interrogate subtle patterns in mutation burden, chromosomal instability, and gene-level alterations across 2,517 H/L and NHW patients. Several key themes emerge: (i) EOCRC remains disproportionately common among H/L individuals, (ii) WNT pathway disruption is nearly ubiquitous across all strata but is modulated by therapy and age, and (iii) the prognostic impact of WNT alterations appears to be context- and ancestry-dependent, particularly in patients who did not receive Bevacizumab.

Clinically, our data reinforce and extend prior observations of an elevated EOCRC burden in H/L populations. H/L patients showed a higher proportion of early-onset disease than NHW patients despite broadly similar sex distribution and stage at diagnosis. The predominance of “colorectal adenocarcinoma NOS” in H/L patients, in contrast to the more granular colon vs. rectal classifications in NHW patients, also raises important questions about diagnostic coding practices, tumor localization, and care fragmentation in underrepresented groups. Together, these patterns underscore the need for ancestry-aware screening strategies and more consistent documentation of tumor site and histology in H/L communities.

At the genomic level, our integrated oncoplot and gene-level analyses confirm that WNT signaling is a central axis of CRC biology across age and ancestry. APC truncating mutations were the dominant event in every subgroup, with WNT pathway alterations present in >80% of tumors regardless of Bevacizumab exposure, age of onset, or ancestry. This pervasive APC-centered disruption supports the concept of WNT activation as a “common trunk” event in colorectal tumorigenesis. However, variation in secondary WNT regulators—RNF43, AXIN1, AXIN2, TCF7L2, AMER1, and CTNNB1, suggests that the route by which tumors amplify or maintain WNT signaling may differ by ancestry, age, and treatment history.

Bevacizumab exposure was consistently associated with lower mutation burden and reduced TMB in both H/L and NHW cohorts, particularly in EOCRC. This pattern, alongside the reduced prevalence of RNF43, AXIN1/2, and TCF7L2 mutations in treated tumors, is compatible with the hypothesis that anti-angiogenic therapy exerts a selection pressure that disfavors certain WNT-altered subclones, or that tumors with less complex WNT mutational architecture are more likely to receive and/or benefit from Bevacizumab. In H/L EOCRC, RNF43 alterations were absent in treated tumors but present in 14.3% of untreated cases, and a similar enrichment of RNF43 and other non-canonical WNT regulators was observed in untreated NHW tumors. These convergent signals identify RNF43 and related genes as potential modulators of Bevacizumab response and motivate functional studies to determine whether specific WNT genotypes confer differential sensitivity to anti-angiogenic therapy.

Age- and ancestry-specific comparisons further refine this picture. Among untreated NHW patients, EOCRC showed a stronger dependence on APC mutations, whereas LOCRC demonstrated enrichment of non-canonical regulators such as AXIN1, AXIN2, and RNF43, suggesting a shift in the architecture of WNT activation with aging. In untreated H/L patients, EOCRC was similarly characterized by a higher APC mutation frequency relative to LOCRC, but subtle increases in CTNNB1 and RNF43 in younger patients hint at additional non-canonical contributions. Cross-ancestry analyses revealed largely overlapping WNT mutational landscapes in Bevacizumab-treated patients, indicating that once therapy is applied, the surviving WNT architecture is remarkably similar between H/L and NHW EOCRC and LOCRC. The one consistent ancestry-related feature among treated EOCRC cases was higher fraction genome altered in H/L tumors compared with NHW, pointing to increased chromosomal instability despite comparable point mutation loads.

Survival analyses revealed that the prognostic meaning of WNT pathway status is not uniform across populations or treatment settings. In H/L patients, both early- and late-onset, treated and untreated, WNT alterations were largely prognostically neutral, with overlapping survival curves and wide confidence intervals, especially in the smaller WNT-unaltered subgroups. In contrast, among NHW patients who did not receive Bevacizumab, WNT-altered tumors were associated with significantly better overall survival in both EOCRC and LOCRC. These findings suggest that, in the absence of anti-angiogenic therapy, WNT-mutated tumors in NHW patients may represent a more “classical” APC-driven biology that remains responsive to standard treatment paradigms, whereas WNT-unaltered tumors may engage alternative pathways with poorer outcomes. The lack of a similar signal in H/L cohorts could reflect smaller sample sizes, differences in comorbidities and access to care, or genuine biological divergence. Importantly, once Bevacizumab was introduced, WNT status ceased to have a detectable survival impact in both ancestries, implying that anti-angiogenic therapy may partially override baseline WNT-associated prognostic patterns or that selected post-Bevacizumab populations are more homogeneous with respect to WNT signaling.

These observations have several implications for precision oncology and health equity. First, the near-universal presence of WNT dysregulation argues against using WNT pathway alteration as a simple binary biomarker in CRC; instead, attention should shift toward specific WNT genotypes (e.g., RNF43-mutant vs. wild-type) and their interaction with therapies such as Bevacizumab. Second, the distinct age- and ancestry-associated patterns, higher chromosomal instability in treated H/L EOCRC, enrichment of non-canonical WNT regulators in older untreated NHW patients, and favorable survival linked to WNT alterations only in NHW cohorts, highlight the danger of extrapolating biomarker findings from one population to another without careful validation. Third, the strong selection signals against certain WNT alterations in Bevacizumab-treated tumors raise the prospect that WNT pathway status could help refine patient selection or guide the development of rational combination regimens that pair anti-angiogenic agents with WNT-targeted strategies.

The AI-HOPE-WNT agent played a central role in enabling these insights by harmonizing large, heterogeneous datasets and allowing rapid, conversational exploration of pathway-level hypotheses across clinically defined subgroups. Rather than replacing traditional statistical pipelines, AI-HOPE-WNT augments them by providing an interactive layer that can iteratively query, visualize, and reinterpret genomic and clinical data in the context of ancestry, age, and treatment. This approach is particularly valuable for health disparities research, where complex interactions between biology, therapy, and structural inequities must be interrogated across multiple strata that are often underpowered in single datasets.

Several limitations must be acknowledged. The analysis relied on retrospective, publicly available cohorts, which may contain incomplete treatment annotations, limited information on dosing and line of therapy, and sparse data on comorbidities, socioeconomic status, and other social determinants of health that likely shape both therapy receipt and outcomes. Some key subgroups, particularly WNT-unaltered H/L patients, were small, leading to wide confidence intervals and reduced power to detect modest survival effects. Our focus on Bevacizumab also does not capture potential interactions between WNT status and other systemic therapies, including targeted agents and immunotherapies. Finally, while AI-HOPE-WNT facilitates hypothesis generation and stratified analyses, causal inferences regarding treatment selection and clonal evolution will require prospective studies with longitudinal sampling and functional validation.

Despite these constraints, our AI-enabled, ancestry-aware evaluation of WNT signaling in CRC provides a nuanced view of how a “ubiquitous” pathway can have context-dependent clinical relevance. By demonstrating that Bevacizumab exposure, age of onset, and ancestry each shape the WNT mutational landscape and its association with survival, this work lays a foundation for future studies that integrate multi-omics, treatment histories, and social context to design more equitable and biologically informed strategies for EOCRC and LOCRC across diverse populations.

## 5. Conclusions

In summary, our analysis reveals that although WNT pathway disruption is a universal feature of colorectal cancer, the patterns and implications of these alterations differ meaningfully by ancestry, age of onset, and Bevacizumab exposure. Early-and late-onset tumors across both H/L and NHW patients share a core dependence on APC inactivation, yet secondary WNT regulators such as RNF43, AXIN1/2, and TCF7L2 exhibit treatment-and ancestry-specific variation that may signal distinct biological trajectories. The contrasting survival associations—neutral in H/L cohorts but beneficial in untreated NHW groups—underscore that WNT alterations do not confer uniform prognostic value across populations. These findings highlight the importance of integrating ancestry-aware genomic characterization with detailed treatment histories to better resolve the heterogeneity of CRC and to inform more precise, equitable therapeutic strategies, particularly for populations with a disproportionate burden of early-onset disease.

## Data Availability

All data used in the present study is publicly available at https://www.cbioportal.org/ and https://genie.cbioportal.org. The datasets used in our study were aggregated/summary data, and no individual-level data were used. Additional data can be provided upon reasonable request to the authors.

## Supplementary Materials

**Table S1-.**
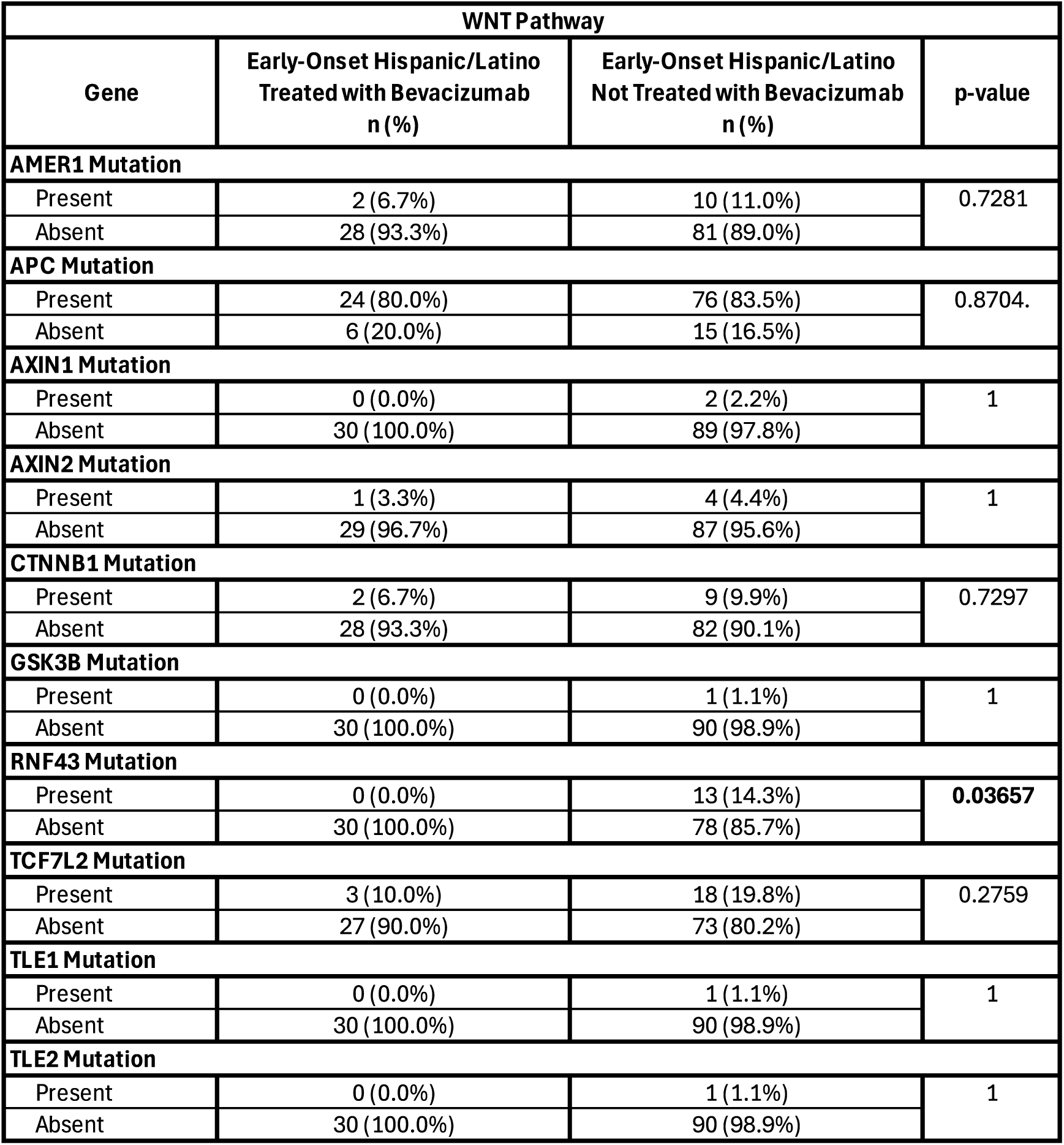
EO HL Treated with Bevacizumab vs EO HL Not Treated with Bevacizumab.

| WNT Pathway |  |  |  |
| --- | --- | --- | --- |
| Gene | Early-Onset Hispanic/Latino<br>Treated with Bevacizumab<br>n (%) | Early-Onset Hispanic/Latino<br>Not Treated with Bevacizumab<br>n (%) | p-value |
| AMER1 Mutation |  |  |  |
| Present | 2 (6.7%) | 10 (11.0%) | 0.7281 |
| Absent | 28 (93.3%) | 81 (89.0%) |  |
| APC Mutation |  |  |  |
| Present | 24 (80.0%) | 76 (83.5%) | 0.8704. |
| Absent | 6 (20.0%) | 15 (16.5%) |  |
| AXIN1 Mutation |  |  |  |
| Present | 0 (0.0%) | 2 (2.2%) | 1 |
| Absent | 30 (100.0%) | 89 (97.8%) |  |
| AXIN2 Mutation |  |  |  |
| Present | 1 (3.3%) | 4 (4.4%) | 1 |
| Absent | 29 (96.7%) | 87 (95.6%) |  |
| CTNNB1 Mutation |  |  |  |
| Present | 2 (6.7%) | 9 (9.9%) | 0.7297 |
| Absent | 28 (93.3%) | 82 (90.1%) |  |
| GSK3B Mutation |  |  |  |
| Present | 0 (0.0%) | 1 (1.1%) | 1 |
| Absent | 30 (100.0%) | 90 (98.9%) |  |
| RNF43 Mutation |  |  |  |
| Present | 0 (0.0%) | 13 (14.3%) | 0.03657 |
| Absent | 30 (100.0%) | 78 (85.7%) |  |
| TCF7L2 Mutation |  |  |  |
| Present | 3 (10.0%) | 18 (19.8%) | 0.2759 |
| Absent | 27 (90.0%) | 73 (80.2%) |  |
| TLE1 Mutation |  |  |  |
| Present | 0 (0.0%) | 1 (1.1%) | 1 |
| Absent | 30 (100.0%) | 90 (98.9%) |  |
| TLE2 Mutation |  |  |  |
| Present | 0 (0.0%) | 1 (1.1%) | 1 |
| Absent | 30 (100.0%) | 90 (98.9%) |  |

**Table S2-.** LO HL Treated with Bevacizumab v LO HL Not Treated with Bevacizumab.

| WNT Pathway |  |  |  |
| --- | --- | --- | --- |
| Gene | Late-Onset Hispanic/Latino<br>Treated with Bevacizumab<br>n (%) | Late-Onset Hispanic/Latino<br>Not Treated with Bevacizumab<br>n (%) | p-value |
| AMER1 Mutation |  |  |  |
| Present | 5 (11.6%) | 7 (6.3%) | 0.4412 |
| Absent | 38 (88.4%) | 104 (93.7%) |  |
| APC Mutation |  |  |  |
| Present | 34 (79.1%) | 73 (65.8%) | 0.1575 |
| Absent | 9 (20.9%) | 38 (34.2%) |  |
| AXIN1 Mutation |  |  |  |
| Present | 0 (0.0%) | 3 (2.7%) | 0.5602 |
| Absent | 43 (100.0%) | 108 (97.3%) |  |
| AXIN2 Mutation |  |  |  |
| Present | 0 (0.0%) | 8 (7.2%) | 0.1067 |
| Absent | 43 (100.0%) | 103 (92.8%) |  |
| CTNNB1 Mutation |  |  |  |
| Present | 3 (7.0%) | 7 (6.3%) | 1 |
| Absent | 40 (93.0%) | 104 (93.7%) |  |
| GSK3B Mutation |  |  |  |
| Present | 0 (0.0%) | 1 (0.9%) | 1 |
| Absent | 43 (100.0%) | 110 (99.1%) |  |
| RNF43 Mutation |  |  |  |
| Present | 1 (2.3%) | 20 (18.0%) | 0.008617 |
| Absent | 42 (97.7%) | 91 (82.0%) |  |
| TCF7L2 Mutation |  |  |  |
| Present | 5 (11.6%) | 15 (13.5%) | 0.964 |
| Absent | 38 (88.4%) | 96 (86.5%) |  |
| TLE1 Mutation |  |  |  |
| Present | 0 (0.0%) | 0 (0.0%) | 1 |
| Absent | 43 (100.0%) | 111 (100.0%) |  |
| TLE2 Mutation |  |  |  |
| Present | 0 (0.0%) | 0 (0.0%) | 1 |
| Absent | 43 (100.0%) | 111 (100.0%) |  |

**Table S3.** EO HL Treated with Bevacizumab v LO HL Treated with Bevacizumab.

| WNT Pathway |  |  |  |
| --- | --- | --- | --- |
| Gene | Early-Onset Hispanic/Latino<br>Treated with Bevacizumab<br>n (%) | Late-Onset Hispanic/Latino<br>Treated with Bevacizumab<br>n (%) | p-value |
| AMER1 Mutation |  |  |  |
| Present | 2 (6.7%) | 5 (11.6%) | 0.6925 |
| Absent | 28 (93.3%) | 38 (88.4%) |  |
| APC Mutation |  |  |  |
| Present | 24 (80.0%) | 34 (79.1%) | 1 |
| Absent | 6 (20.0%) | 9 (20.9%) |  |
| AXIN1 Mutation |  |  |  |
| Present | 0 (0.0%) | 0 (0.0%) | 1 |
| Absent | 30 (100.0%) | 43 (100.0%) |  |
| AXIN2 Mutation |  |  |  |
| Present | 1 (3.3%) | 0 (0.0%) | 0.411 |
| Absent | 29 (96.7%) | 43 (100.0%) |  |
| CTNNB1 Mutation |  |  |  |
| Present | 2 (6.7%) | 3 (7.0%) | 1 |
| Absent | 28 (93.3%) | 40 (93.0%) |  |
| GSK3B Mutation |  |  |  |
| Present | 0 (0.0%) | 0 (0.0%) | 1 |
| Absent | 30 (100.0%) | 43 (100.0%) |  |
| RNF43 Mutation |  |  |  |
| Present | 0 (0.0%) | 1 (2.3%) | 1 |
| Absent | 30 (100.0%) | 42 (97.7%) |  |
| TCF7L2 Mutation |  |  |  |
| Present | 3 (10.0%) | 5 (11.6%) | 1 |
| Absent | 27 (90.0%) | 38 (88.4%) |  |
| TLE1 Mutation |  |  |  |
| Present | 0 (0.0%) | 0 (0.0%) | 1 |
| Absent | 30 (100.0%) | 43 (100.0%) |  |
| TLE2 Mutation |  |  |  |
| Present | 0 (0.0%) | 0 (0.0%) | 1 |
| Absent | 30 (100.0%) | 43 (100.0%) |  |

**Table S4.**
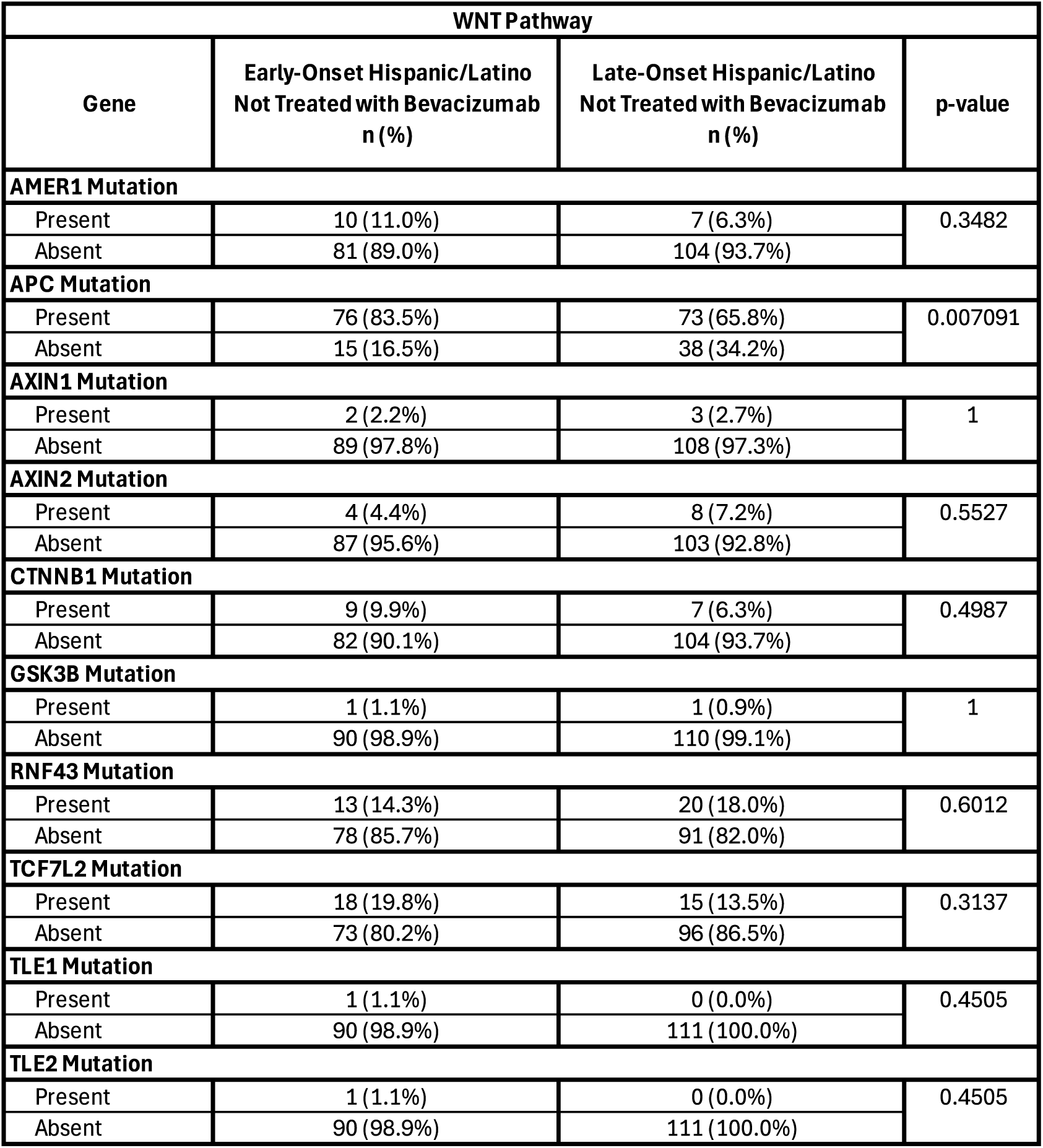
EO HL Not Treated with Bevacizumab v LO HL Not Treated with Bevacizumab.

| WNT Pathway |  |  |  |
| --- | --- | --- | --- |
| Gene | Early-Onset Hispanic/Latino<br>Not Treated with Bevacizumab<br>n (%) | Late-Onset Hispanic/Latino<br>Not Treated with Bevacizumab<br>n (%) | p-value |
| AMER1 Mutation |  |  |  |
| Present | 10 (11.0%) | 7 (6.3%) | 0.3482 |
| Absent | 81 (89.0%) | 104 (93.7%) |  |
| APC Mutation |  |  |  |
| Present | 76 (83.5%) | 73 (65.8%) | 0.007091 |
| Absent | 15 (16.5%) | 38 (34.2%) |  |
| AXIN1 Mutation |  |  |  |
| Present | 2 (2.2%) | 3 (2.7%) | 1 |
| Absent | 89 (97.8%) | 108 (97.3%) |  |
| AXIN2 Mutation |  |  |  |
| Present | 4 (4.4%) | 8 (7.2%) | 0.5527 |
| Absent | 87 (95.6%) | 103 (92.8%) |  |
| CTNNB1 Mutation |  |  |  |
| Present | 9 (9.9%) | 7 (6.3%) | 0.4987 |
| Absent | 82 (90.1%) | 104 (93.7%) |  |
| GSK3B Mutation |  |  |  |
| Present | 1 (1.1%) | 1 (0.9%) | 1 |
| Absent | 90 (98.9%) | 110 (99.1%) |  |
| RNF43 Mutation |  |  |  |
| Present | 13 (14.3%) | 20 (18.0%) | 0.6012 |
| Absent | 78 (85.7%) | 91 (82.0%) |  |
| TCF7L2 Mutation |  |  |  |
| Present | 18 (19.8%) | 15 (13.5%) | 0.3137 |
| Absent | 73 (80.2%) | 96 (86.5%) |  |
| TLE1 Mutation |  |  |  |
| Present | 1 (1.1%) | 0 (0.0%) | 0.4505 |
| Absent | 90 (98.9%) | 111 (100.0%) |  |
| TLE2 Mutation |  |  |  |
| Present | 1 (1.1%) | 0 (0.0%) | 0.4505 |
| Absent | 90 (98.9%) | 111 (100.0%) |  |

**Table S5.** EO NHW Treated with Bevacizumab v EO NHW Not Treated with Bevacizumab.

| WNT Pathway |  |  |  |
| --- | --- | --- | --- |
| Gene | Early-Onset NHW<br>Treated with Bevacizumab<br>n (%) | Early-Onset NHW<br>Not Treated with Bevacizumab<br>n (%) | p-value |
| AMER1 Mutation |  |  |  |
| Present | 8 (4.7%) | 45 (7.9%) | 0.1992 |
| Absent | 164 (95.3%) | 524 (92.1%) |  |
| APC Mutation |  |  |  |
| Present | 129 (75.0%) | 460 (80.8%) | 0.1199 |
| Absent | 43 (25.0%) | 109 (19.2%) |  |
| AXIN1 Mutation |  |  |  |
| Present | 0 (0.0%) | 15 (2.6%) | 0.02845 |
| Absent | 172 (100.0%) | 554 (97.4%) |  |
| AXIN2 Mutation |  |  |  |
| Present | 2 (1.2%) | 29 (5.1%) | 0.02724 |
| Absent | 170 (98.8%) | 540 (94.9%) |  |
| CTNNB1 Mutation |  |  |  |
| Present | 10 (5.8%) | 34 (6.0%) | 1 |
| Absent | 162 (94.2%) | 535 (94.0%) |  |
| GSK3B Mutation |  |  |  |
| Present | 1 (0.6%) | 6 (1.1%) | 1 |
| Absent | 171 (99.4%) | 563 (98.9%) |  |
| RNF43 Mutation |  |  |  |
| Present | 3 (1.7%) | 44 (7.7%) | 0.003625 |
| Absent | 169 (98.3%) | 525 (92.3%) |  |
| TCF7L2 Mutation |  |  |  |
| Present | 20 (11.6%) | 119 (20.9%) | 0.008735 |
| Absent | 152 (88.4%) | 450 (79.1%) |  |
| TLE1 Mutation |  |  |  |
| Present | 0 (0.0%) | 0 (0.0%) | 1 |
| Absent | 172 (100.0%) | 569 (100.0%) |  |
| TLE2 Mutation |  |  |  |
| Present | 0 (0.0%) | 0 (0.0%) | 1 |
| Absent | 172 (100.0%) | 569 (100.0%) |  |

**Table S6.** LO NHW Treated with Bevacizumab v LO NHW Not Treated with Bevacizumab.

| WNT Pathway |  |  |  |
| --- | --- | --- | --- |
| Gene | Late-Onset NHW<br>Treated with Bevacizumab<br>n (%) | Late-Onset NHW<br>Not Treated with Bevacizumab<br>n (%) | p-value |
| AMER1 Mutation |  |  |  |
| Present | 22 (5.9%) | 146 (11.0%) | 0.005384 |
| Absent | 349 (94.1%) | 1184 (89.0%) |  |
| APC Mutation |  |  |  |
| Present | 297 (80.1%) | 999 (75.1%) | 0.05653 |
| Absent | 74 (19.9%) | 331 (24.9%) |  |
| AXIN1 Mutation |  |  |  |
| Present | 3 (0.8%) | 63 (4.7%) | 0.0001839 |
| Absent | 368 (99.2%) | 1267 (95.3%) |  |
| AXIN2 Mutation |  |  |  |
| Present | 14 (3.8%) | 110 (8.3%) | 0.004607 |
| Absent | 357 (96.2%) | 1220 (91.7%) |  |
| CTNNB1 Mutation |  |  |  |
| Present | 22 (5.9%) | 93 (7.0%) | 0.5459 |
| Absent | 349 (94.1%) | 1237 (93.0%) |  |
| GSK3B Mutation |  |  |  |
| Present | 1 (0.3%) | 17 (1.3%) | 0.1465 |
| Absent | 370 (99.7%) | 1313 (98.7%) |  |
| RNF43 Mutation |  |  |  |
| Present | 18 (4.9%) | 183 (13.8%) | 4.05E-06 |
| Absent | 353 (95.1%) | 1147 (86.2%) |  |
| TCF7L2 Mutation |  |  |  |
| Present | 44 (11.9%) | 225 (16.9%) | 0.01396 |
| Absent | 327 (88.1%) | 1105 (83.1%) |  |
| TLE1 Mutation |  |  |  |
| Present | 0 (0.0%) | 0 (0.0%) | 1 |
| Absent | 371 (100.0%) | 1330 (100.0%) |  |
| TLE2 Mutation |  |  |  |
| Present | 0 (0.0%) | 0 (0.0%) | 1 |
| Absent | 371 (100.0%) | 1330 (100.0%) |  |

**Table S7.**
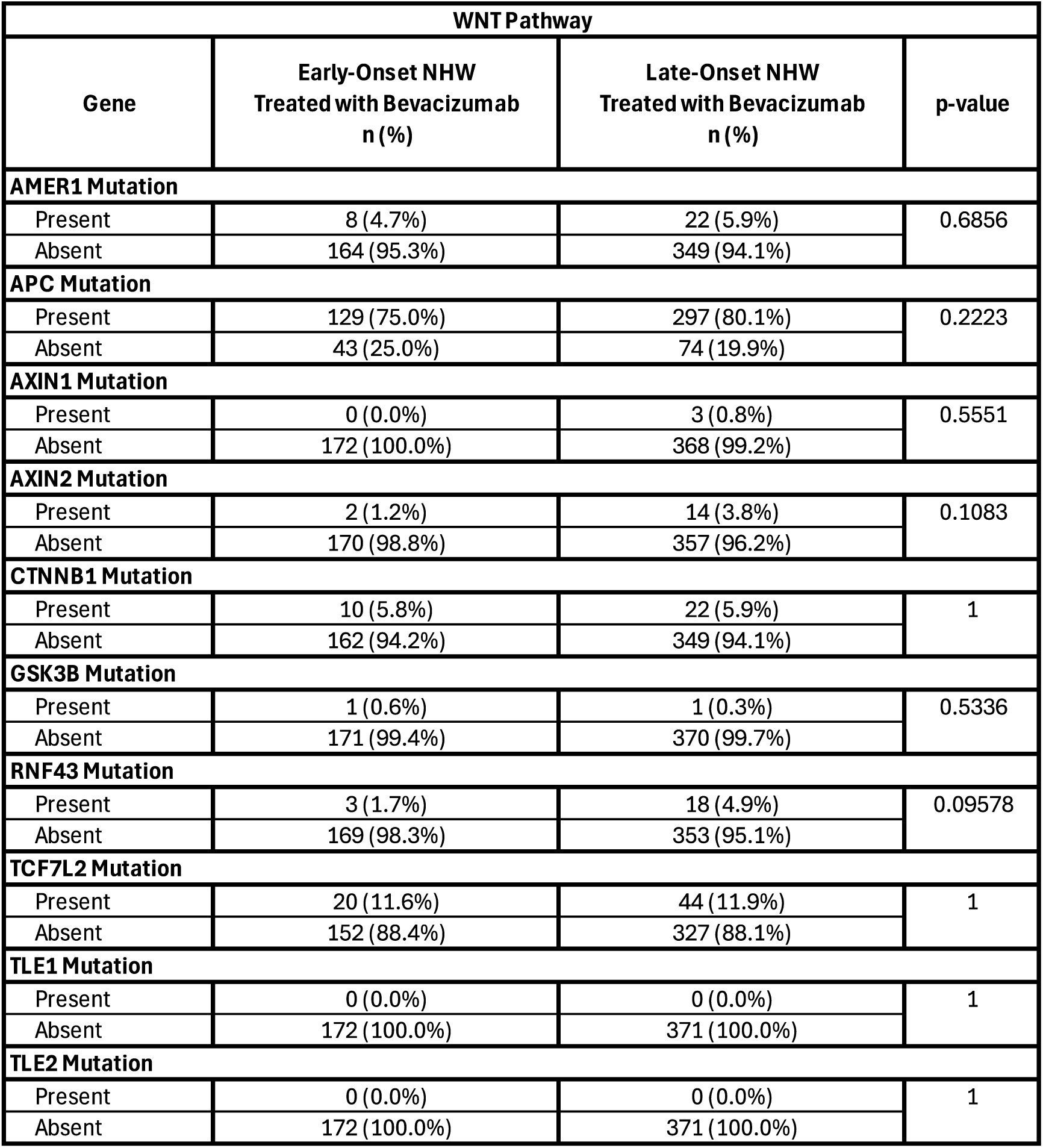
EO NHW Treated with Bevacizumab v LO NHW Treated with Bevacizumab.

**Table S8.** EO NHW Not Treated with Bevacizumab v LO NHW Not Treated with Bevacizumab.

| WNT Pathway |  |  |  |
| --- | --- | --- | --- |
| Gene | Early-Onset NHW<br>Not Treated with Bevacizumab<br>n (%) | Late-Onset NHW<br>Not Treated with Bevacizumab<br>n (%) | p-value |
| AMER1 Mutation |  |  |  |
| Present | 45 (7.9%) | 146 (11.0%) | 0.05075 |
| Absent | 524 (92.1%) | 1184 (89.0%) |  |
| APC Mutation |  |  |  |
| Present | 460 (80.8%) | 999 (75.1%) | 0.007999 |
| Absent | 109 (19.2%) | 331 (24.9%) |  |
| AXIN1 Mutation |  |  |  |
| Present | 15 (2.6%) | 63 (4.7%) | 0.04695 |
| Absent | 554 (97.4%) | 1267 (95.3%) |  |
| AXIN2 Mutation |  |  |  |
| Present | 29 (5.1%) | 110 (8.3%) | 0.01946 |
| Absent | 540 (94.9%) | 1220 (91.7%) |  |
| CTNNB1 Mutation |  |  |  |
| Present | 34 (6.0%) | 93 (7.0%) | 0.4762 |
| Absent | 535 (94.0%) | 1237 (93.0%) |  |
| GSK3B Mutation |  |  |  |
| Present | 6 (1.1%) | 17 (1.3%) | 0.8577 |
| Absent | 563 (98.9%) | 1313 (98.7%) |  |
| RNF43 Mutation |  |  |  |
| Present | 44 (7.7%) | 183 (13.8%) | 0.0002822 |
| Absent | 525 (92.3%) | 1147 (86.2%) |  |
| TCF7L2 Mutation |  |  |  |
| Present | 119 (20.9%) | 225 (16.9%) | 0.0448 |
| Absent | 450 (79.1%) | 1105 (83.1%) |  |
| TLE1 Mutation |  |  |  |
| Present | 0 (0.0%) | 0 (0.0%) | 1 |
| Absent | 569 (100.0%) | 1330 (100.0%) |  |
| TLE2 Mutation |  |  |  |
| Present | 0 (0.0%) | 0 (0.0%) | 1 |
| Absent | 569 (100.0%) | 1330 (100.0%) |  |

**Table S9.** EO HL Treated with Bevacizumab v EO NHW Treated with Bevacizumab.

| WNT Pathway |  |  |  |
| --- | --- | --- | --- |
| Gene | Early-Onset Hispanic/Latino<br>Treated with Bevacizumab<br>n (%) | Early-Onset NHW<br>Treated with Bevacizumab<br>n (%) | p-value |
| AMER1 Mutation |  |  |  |
| Present | 2 (6.7%) | 8 (4.7%) | 0.6458 |
| Absent | 28 (93.3%) | 164 (95.3%) |  |
| APC Mutation |  |  |  |
| Present | 24 (80.0%) | 129 (75.0%) | 0.7198 |
| Absent | 6 (20.0%) | 43 (25.0%) |  |
| AXIN1 Mutation |  |  |  |
| Present | 0 (0.0%) | 0 (0.0%) | 1 |
| Absent | 30 (100.0%) | 172 (100.0%) |  |
| AXIN2 Mutation |  |  |  |
| Present | 1 (3.3%) | 2 (1.2%) | 0.3843 |
| Absent | 29 (96.7%) | 170 (98.8%) |  |
| CTNNB1 Mutation |  |  |  |
| Present | 2 (6.7%) | 10 (5.8%) | 0.694 |
| Absent | 28 (93.3%) | 162 (94.2%) |  |
| GSK3B Mutation |  |  |  |
| Present | 0 (0.0%) | 1 (0.6%) | 1 |
| Absent | 30 (100.0%) | 171 (99.4%) |  |
| RNF43 Mutation |  |  |  |
| Present | 0 (0.0%) | 3 (1.7%) | 1 |
| Absent | 30 (100.0%) | 169 (98.3%) |  |
| TCF7L2 Mutation |  |  |  |
| Present | 3 (10.0%) | 20 (11.6%) | 1 |
| Absent | 27 (90.0%) | 152 (88.4%) |  |
| TLE1 Mutation |  |  |  |
| Present | 0 (0.0%) | 0 (0.0%) | 1 |
| Absent | 30 (100.0%) | 172 (100.0%) |  |
| TLE2 Mutation |  |  |  |
| Present | 0 (0.0%) | 0 (0.0%) | 1 |
| Absent | 30 (100.0%) | 172 (100.0%) |  |

**Table S10.** EO HL Not Treated with Bevacizumab v EO NHW Not Treated with Bevacizumab.

| WNT Pathway |  |  |  |
| --- | --- | --- | --- |
| Gene | Early-Onset Hispanic/Latino<br>Not Treated with Bevacizumab<br>n (%) | Early-Onset NHW<br>Not Treated with Bevacizumab<br>n (%) | p-value |
| AMER1 Mutation |  |  |  |
| Present | 10 (11.0%) | 45 (7.9%) | 0.4337 |
| Absent | 81 (89.0%) | 524 (92.1%) |  |
| APC Mutation |  |  |  |
| Present | 76 (83.5%) | 460 (80.8%) | 0.6444 |
| Absent | 15 (16.5%) | 109 (19.2%) |  |
| AXIN1 Mutation |  |  |  |
| Present | 2 (2.2%) | 15 (2.6%) | 1 |
| Absent | 89 (97.8%) | 554 (97.4%) |  |
| AXIN2 Mutation |  |  |  |
| Present | 4 (4.4%) | 29 (5.1%) | 1 |
| Absent | 87 (95.6%) | 540 (94.9%) |  |
| CTNNB1 Mutation |  |  |  |
| Present | 9 (9.9%) | 34 (6.0%) | 0.2395 |
| Absent | 82 (90.1%) | 535 (94.0%) |  |
| GSK3B Mutation |  |  |  |
| Present | 1 (1.1%) | 6 (1.1%) | 1 |
| Absent | 90 (98.9%) | 563 (98.9%) |  |
| RNF43 Mutation |  |  |  |
| Present | 13 (14.3%) | 44 (7.7%) | 0.06214 |
| Absent | 78 (85.7%) | 525 (92.3%) |  |
| TCF7L2 Mutation |  |  |  |
| Present | 18 (19.8%) | 119 (20.9%) | 0.9137 |
| Absent | 73 (80.2%) | 450 (79.1%) |  |
| TLE1 Mutation |  |  |  |
| Present | 1 (1.1%) | 0 (0.0%) | 0.1379 |
| Absent | 90 (98.9%) | 569 (100.0%) |  |
| TLE2 Mutation |  |  |  |
| Present | 1 (1.1%) | 0 (0.0%) | 0.1379 |
| Absent | 90 (98.9%) | 569 (100.0%) |  |

**Table S11.** LO HL Treated with Bevacizumab v LO NHW Treated with Bevacizumab.

| WNT Pathway |  |  |  |
| --- | --- | --- | --- |
| Gene | Late-Onset Hispanic/Latino<br>Treated with Bevacizumab<br>n (%) | Late-Onset NHW<br>Treated with Bevacizumab<br>n (%) | p-value |
| AMER1 Mutation |  |  |  |
| Present | 5 (11.6%) | 22 (5.9%) | 0.2686 |
| Absent | 38 (88.4%) | 349 (94.1%) |  |
| APC Mutation |  |  |  |
| Present | 34 (79.1%) | 297 (80.1%) | 1 |
| Absent | 9 (20.9%) | 74 (19.9%) |  |
| AXIN1 Mutation |  |  |  |
| Present | 0 (0.0%) | 3 (0.8%) | 1 |
| Absent | 43 (100.0%) | 368 (99.2%) |  |
| AXIN2 Mutation |  |  |  |
| Present | 0 (0.0%) | 14 (3.8%) | 0.3788 |
| Absent | 43 (100.0%) | 357 (96.2%) |  |
| CTNNB1 Mutation |  |  |  |
| Present | 3 (7.0%) | 22 (5.9%) | 0.735 |
| Absent | 40 (93.0%) | 349 (94.1%) |  |
| GSK3B Mutation |  |  |  |
| Present | 0 (0.0%) | 1 (0.3%) | 1 |
| Absent | 43 (100.0%) | 370 (99.7%) |  |
| RNF43 Mutation |  |  |  |
| Present | 1 (2.3%) | 18 (4.9%) | 0.7072 |
| Absent | 42 (97.7%) | 353 (95.1%) |  |
| TCF7L2 Mutation |  |  |  |
| Present | 5 (11.6%) | 44 (11.9%) | 1 |
| Absent | 38 (88.4%) | 327 (88.1%) |  |
| TLE1 Mutation |  |  |  |
| Present | 0 (0.0%) | 0 (0.0%) | 1 |
| Absent | 43 (100.0%) | 371 (100.0%) |  |
| TLE2 Mutation |  |  |  |
| Present | 0 (0.0%) | 0 (0.0%) | 1 |
| Absent | 43 (100.0%) | 371 (100.0%) |  |

**Table S12.** LO HL Not Treated with Bevacizumab v LO NHW Not Treated with Bevacizumab.

| WNT Pathway |  |  |  |
| --- | --- | --- | --- |
| Gene | Late-Onset Hispanic/Latino<br>Not Treated with Bevacizumab<br>n (%) | Late-Onset NHW<br>Not Treated with Bevacizumab<br>n (%) | p-value |
| AMER1 Mutation |  |  |  |
| Present | 7 (6.3%) | 146 (11.0%) | 0.1693 |
| Absent | 104 (93.7%) | 1184 (89.0%) |  |
| APC Mutation |  |  |  |
| Present | 73 (65.8%) | 999 (75.1%) | 0.03993 |
| Absent | 38 (34.2%) | 331 (24.9%) |  |
| AXIN1 Mutation |  |  |  |
| Present | 3 (2.7%) | 63 (4.7%) | 0.4766 |
| Absent | 108 (97.3%) | 1267 (95.3%) |  |
| AXIN2 Mutation |  |  |  |
| Present | 8 (7.2%) | 110 (8.3%) | 0.8318 |
| Absent | 103 (92.8%) | 1220 (91.7%) |  |
| CTNNB1 Mutation |  |  |  |
| Present | 7 (6.3%) | 93 (7.0%) | 0.9371 |
| Absent | 104 (93.7%) | 1237 (93.0%) |  |
| GSK3B Mutation |  |  |  |
| Present | 1 (0.9%) | 17 (1.3%) | 1 |
| Absent | 110 (99.1%) | 1313 (98.7%) |  |
| RNF43 Mutation |  |  |  |
| Present | 20 (18.0%) | 183 (13.8%) | 0.2726 |
| Absent | 91 (82.0%) | 1147 (86.2%) |  |
| TCF7L2 Mutation |  |  |  |
| Present | 15 (13.5%) | 225 (16.9%) | 0.4283 |
| Absent | 96 (86.5%) | 1105 (83.1%) |  |
| TLE1 Mutation |  |  |  |
| Present | 0 (0.0%) | 0 (0.0%) | 1 |
| Absent | 111 (100.0%) | 1330 (100.0%) |  |
| TLE2 Mutation |  |  |  |
| Present | 0 (0.0%) | 0 (0.0%) | 1 |
| Absent | 111 (100.0%) | 1330 (100.0%) |  |

**Table S13.**
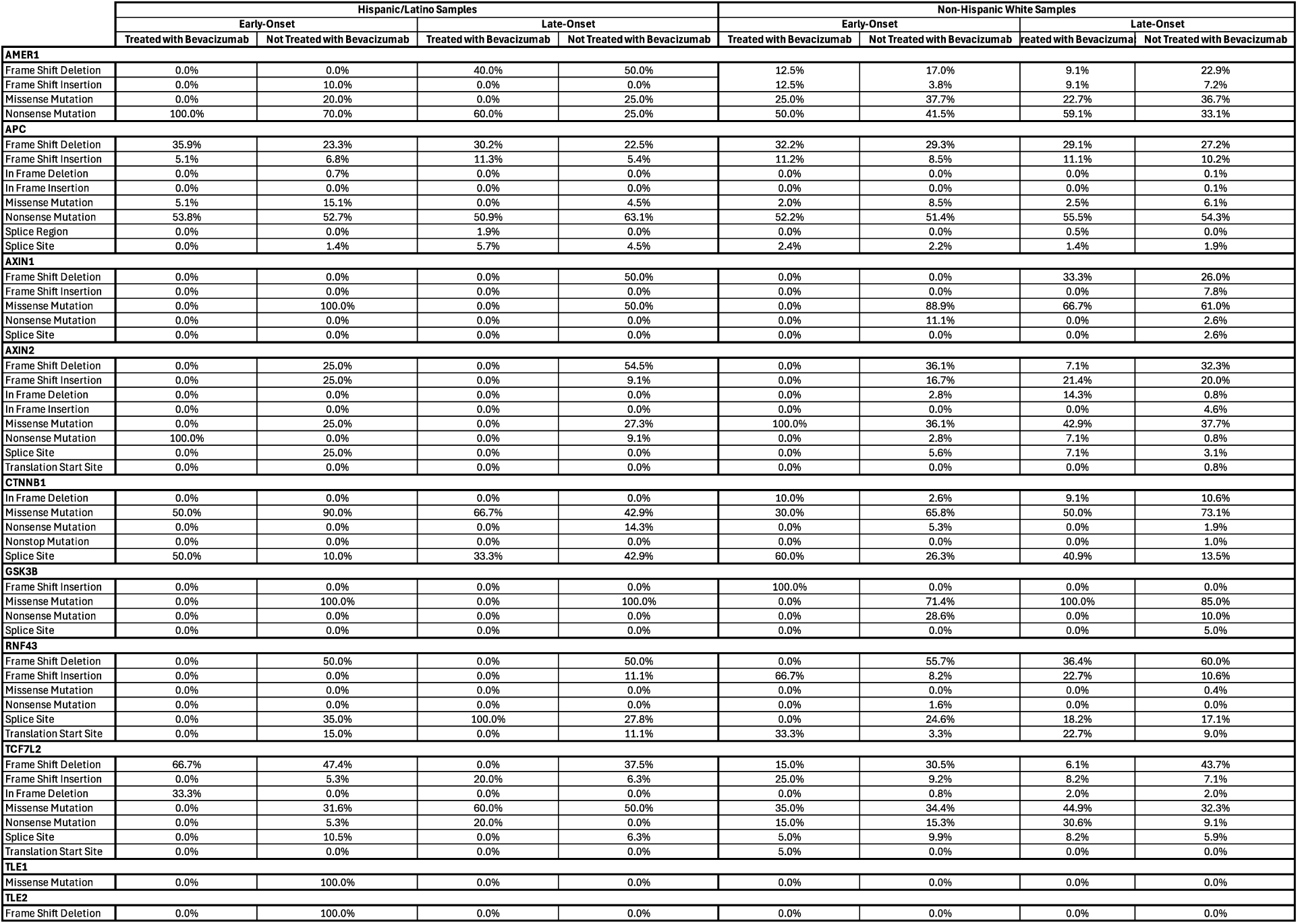
Mutational landscape of H/L and NHW early- and late-onset CRC with and without Bevacizumab treatment.

**Figure S1.**
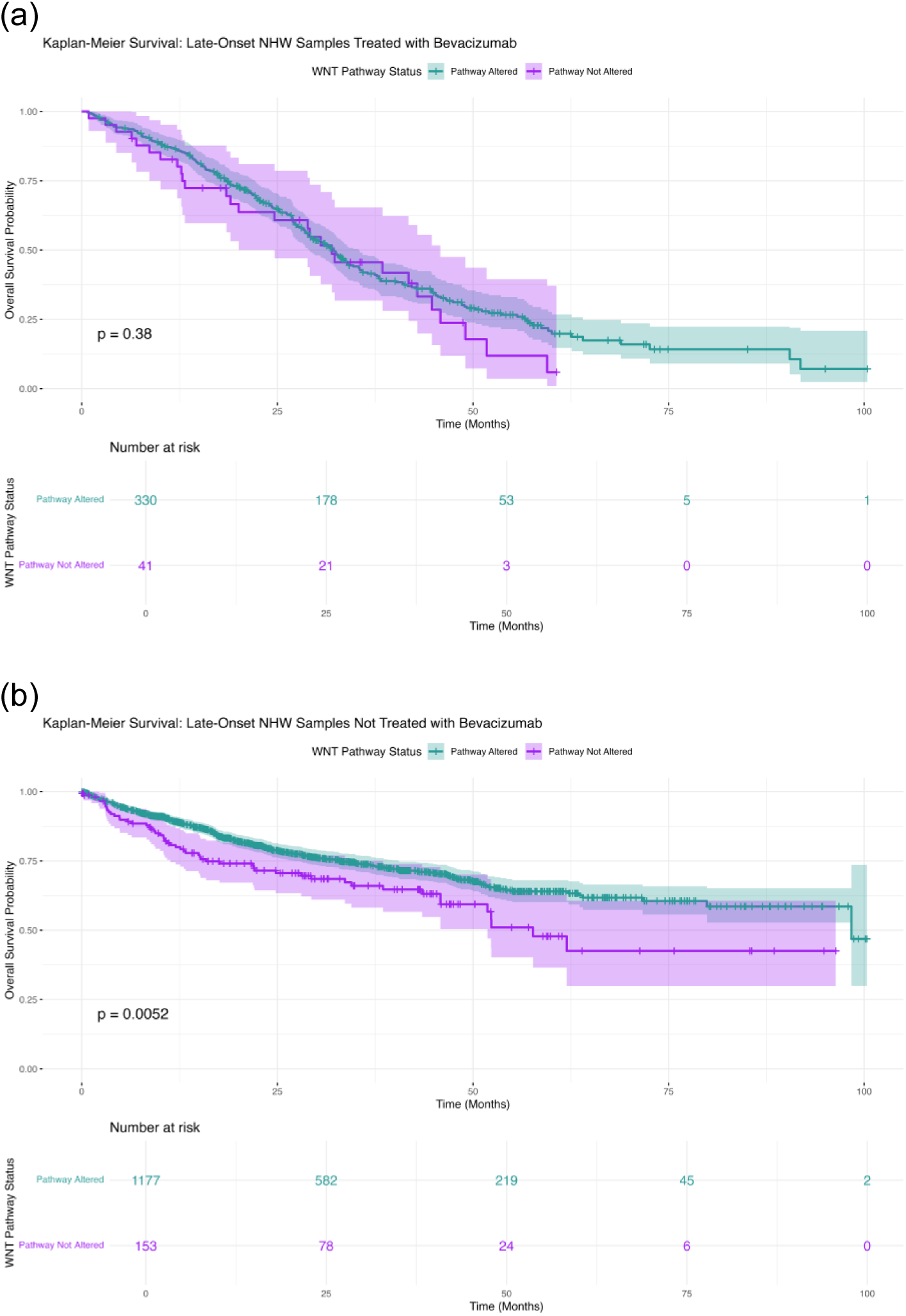
Somatic mutation profiles of WNT pathway genes stratified by age, ethnicity, and Bevacizumab treatment status in colorectal cancer. Oncoplot visualizations depict the distribution and frequency of mutation types across WNT pathway genes for two patient groups: (a) Late-Onset NHW patients treated with Bevacizumab, and (b) Late-Onset NHW patients not receiving Bevacizumab.

